# Single-cell sequencing of the human midbrain reveals glial activation and a neuronal state specific to Parkinson’s disease

**DOI:** 10.1101/2020.09.28.20202812

**Authors:** S. Smajić, C. A. Prada-Medina, Z. Landoulsi, C. Dietrich, J. Jarazo, J. Henck, S. Balachandran, S. Pachchek, C. M. Morris, P. Antony, B. Timmermann, S. Sauer, J. C. Schwamborn, P. May, A. Grünewald, M. Spielmann

## Abstract

Parkinson’s disease (PD) etiology is associated with genetic and environmental factors that lead to a loss of dopaminergic neurons. However, the functional interpretation of PD-associated risk variants and how other midbrain cells contribute to this neurodegenerative process are poorly understood. Here, we profiled >41,000 single-nuclei transcriptomes of postmortem midbrain tissue from 6 idiopathic PD (IPD) patients and 5 matched controls. We show that PD-risk variants are associated with glia- and neuron-specific gene expression patterns. Furthermore, Microglia and astrocytes presented IPD-specific cell proliferation and dysregulation of genes related to unfolded protein response and cytokine signalling. IPD-microglia revealed a specific pro-inflammatory trajectory. Finally, we discovered a neuronal cell cluster exclusively present in IPD midbrains characterized by *CADPS2* overexpression and a high proportion of cycling cells. We conclude that elevated CADPS2 expression is specific to dysfunctional dopaminergic neurons, which have lost their dopaminergic identity and unsuccessful attempt to re-enter the cell cycle.

## Introduction

Parkinson’s disease (PD) is a neurological disorder that is characterized by a progressive loss of neuromelanin-containing dopaminergic neurons (DaNs) in the substantia nigra (SN)^1,2^. Age, genetic, and environmental factors contribute to PD pathogenesis, but disease pathology and etiology remain mostly unknown^3^. Approximately 95% of PD patients do not harbor an interpretable genetic cause; therefore, they are classified as idiopathic PD (IPD)^4^. This lack of knowledge about the molecular mechanisms underlying neurodegeneration in PD represents a major challenge for developing effective therapies^5^.

Our current understanding of IPD cellular and molecular pathophysiology largely relies on experimental models that lack adequate representation of the disease complexity. For instance, toxin-induced animal models neither capture the human brain’s nature nor the multifactorial aspect of the disease^6^. Also, IPD-patient derived pluripotent stem cell (iPSC) models recapitulate molecular mechanisms underlying IPD pathogenesis but lack the cellular composition dynamics found in the human brain. Moreover, several transcriptomic studies using human postmortem midbrain tissue have investigated the transcriptional programs disrupted in IPD. Most of these studies used bulk RNA-seq approaches, which fail to dissect cell-type-specific contributions to disease pathology^7^. The recent development of single-cell sequencing technologies offers the possibility to overcome these challenges. In particular, transcriptional profiling of single cells (scRNA-seq) or nuclei (snRNA-seq) has proved to be an effective strategy to obtain a global view of disease-associated changes at an unprecedented resolution^8^.

In this study, we performed snRNA-seq of postmortem adult human midbrain tissue of IPD patients and age-matched healthy controls to obtain an unbiased and global view of the cell-type composition and cellular phenotype of IPD at single-cell resolution.

## Results

We sampled adult human postmortem midbrain from five IPD cases, for which pathology reports describe a severe neuronal loss in the SN and no family history of PD (**table S1**). We confirmed the PD cases’ idiopathic nature by SNP-Chip profiling of 179.467 known variants associated with neurological diseases, including PD^9^, which did not reveal a genetic etiology (**table S2**). We sampled six control midbrain to match the IPD patient characteristics. The average age of IPD patients and healthy control individuals were ∼77 and ∼81 years, respectively, and both groups had similar postmortem intervals, IPD ∼22, and controls ∼16 hours (**table S1**).

We sequenced single nuclei from frozen ventral sections of human postmortem midbrains (**Fig. 1A**). Using six to eight sections of each individual’s left hemi-midbrain, we prepared single nuclei suspension by scraping off the tissue from the glass slides and using a modified version of the standard 10X Genomics nuclei isolation protocol (see methods). Single-nuclei barcoded cDNA library was prepared using the 10X Genomics Chromium System and sequenced on the NovaSeq 6000 Illumina platform following the manufacturer recommended protocols. We obtained ∼2000-6000 high-quality nuclei per sample with an average of ∼7,600 transcripts and ∼2,700 genes per nucleus after filtering out poorly sequenced nuclei and potential duplets (**Fig. 1B**). They comprise 22,433 and 19,002 single nuclei from control individuals and IPD patients, respectively (**Fig. 1C**).

**Fig 1.**
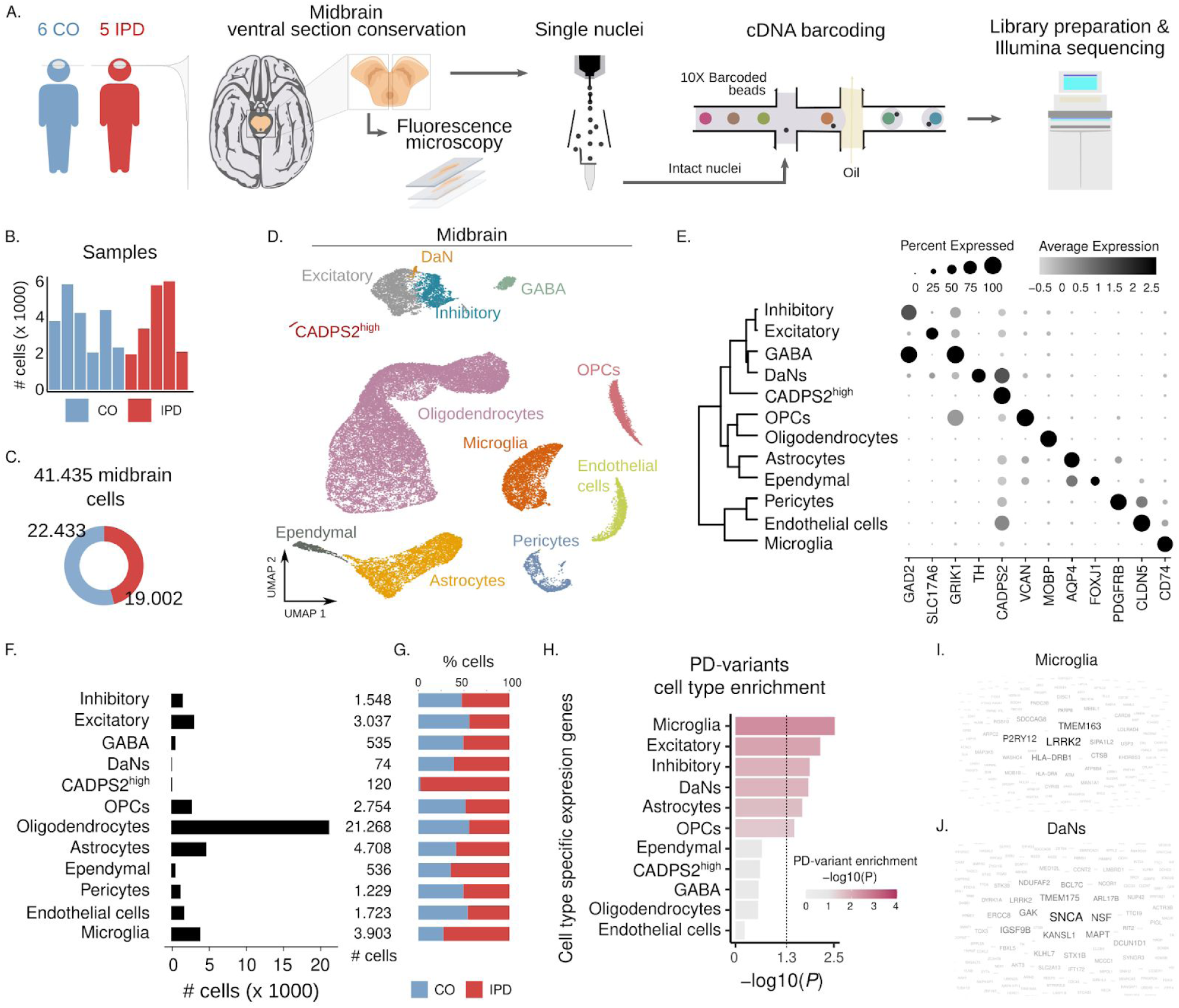
Cell type composition of human midbrain. (**A**) Experimental approach to midbrain tissue processing and nuclei extraction. Nuclei suspensions were processed with the 10x Genomics platform and sequenced with an Illumina sequencer. (**B**) UMAP embedding of the 41,435 human midbrain nuclei. Cells are colored by cell type. (**C**) Number of high-quality obtained nuclei per sample. Overall, the population consists of 19,002 nuclei from IPD patients and 22,433 nuclei from controls. (**D**) Contribution of nuclei from IPD patients or controls to each cell type. (**E**) Cell type transcriptome similarity and representative marker genes. CADPS2^high^ cells cluster together with the neuronal cells. (**F**) Number of profiled nuclei per cell type. (**G**) Proportion of IPD and control profiled cells per cell type. (**H**) PD-associated genomic variants enriched cell type-specific genes. PD-associated variant enrichment (MAGMA) in genes with cell type-specific expression patterns. (**I-J**) Enriched cell type-specific genes of microglia (**I**) and DaNs (**J**) PD genetically associated midbrain cell types. Gene size is proportional to the gene association for a given cell-type within the MAGMA enrichment analysis. This approach associates PD risk variants with cell-type specific expression patterns.

We embedded the 41,435 nuclei transcriptomes into two dimensions using the uniform manifold approximation and projection (UMAP) algorithm. UMAP plots are shown before and after batch correction (**fig S1A, B**). We found that this overall cluster structure was mostly driven by cell-type identity and the inter-sample variability. Interestingly, patient and control cells segregated together within the main clusters (**fig. S1B**). To account for this inter-individual variation for cell-type identification, we followed the Seurat3 sample integration protocol (see methods) (**Fig. 1D**). Using this corrected PCA embedding and the unsupervised and network-based Louvain clustering approach, we found that the human midbrain comprises 12 major cell types (**Fig. 1D, E, fig S1 C**).

The human midbrain is composed of glial, neuronal, vascular, and microglia cells (**Fig. 1D, E**). We annotated most cell clusters by manually comparing well-known marker genes in the literature and the identified marker genes of each cell cluster (**fig. S1C, table S3**). Oligodendrocytes, the most abundant cell-type in the midbrain (**Fig. 1F**), are characterized by the expression of *MOBP*^*10*^, oligodendrocyte precursors cells (OPCs) highly express *VCAN*^*11*^. Expression of *AQP4* is characteristic of astrocytes^12^ and *FOXJ1 of* ependymal cells^13^ (**Fig. 1E-G, fig. S1C**). Also, immune and vascular cells displayed a highly specific expression of well-known marker genes. *CD74* in microglia^14^, *CLDN5* in endothelial cells^15,16^, and *PDGFRB* in pericytes^17^ (**Fig. 1E-G, fig. S1C**). Regarding neuronal cells, we identified four cell-types, excitatory (*SLC17A6*)^18^, inhibitory (*GAD2*)^18^, GABAergic (*GAD2*/*GRIK1*)^19,20^ and, dopaminergic neurons (*TH*)^21^ **(Fig. 1E-G, fig. S1C)**. We also discovered a neuronal cluster of 120 cells, which we could not annotate, characterized by high expression of *CADPS2* (CADPS2^high^ cells) (**Fig. 1E-F, fig. S1C**). These cells almost exclusively originated from IPD patients (IPD, 98.4%; control, 1.6%) **(Fig. 1G)**.

To gain mechanistic insight into genetic risk factors for PD, we evaluated if PD-associated variants are enriched in genomic regions with genes whose expression pattern is cell-type-specific. We found that PD risk variants are associated with microglia and neuronal-specific genes (dopaminergic, excitatory, and inhibitory) (**Fig. 1H, table S4**). They are less associated with astrocytes and OPCs cell-type-specific genes (**Fig. 1H, table S4**). We prioritized the cell-type-specific and PD-risk associated genes based on their enrichment contribution for each cell-type. For instance, we found that LRRK2 showed the highest association with microglia (**Fig. 1I, table S5**) and SNCA was the most prominent PD-associated gene in DaNs (**Fig. 1J**). These findings are in line with previous reports, which identified that PD-associated mutations in alpha-synuclein promote Lewy bodies formation in DaNs^22^ and supports the role of LRRK2 mutations in the activation of microglia in PD^23^. These results, therefore, constitute a valuable resource to prioritize PD-associated variants functionally.

To investigate the cell-type composition changes of the midbrain associated with IPD, we followed three approaches. We compared IPD and control cell density distributions in the two-dimensional UMAP representation (**Fig. 2A-B**) and compared the IPD and control distributions of the cell-type proportions per sample (**Fig. 2C, 2F, 4A, and S3**). Altogether, these results revealed an increase in the fraction of microglia and astrocytes and a decreased fraction of oligodendrocytes in IPD midbrains compared to controls (**Fig. 2, S3, and table S6**). Performing multi-labeling immunofluorescence analysis^24^, we confirmed the increased fraction of microglia and astrocytes in IPD midbrains. We examined paraformaldehyde-fixed paraffin-embedded sections from the right hemi-midbrain of the age and gender-matched pair C3 and IPD3 (**cp. table S1**). We labeled microglia and astrocytes with antibodies against their marker proteins IBA1 and GFAP, respectively (**Fig. 2D, G**). Automated image analysis confirmed an increase in microglia and astrocyte areas as well as in IBA1 and GFAP protein abundance in IPD3 compared to C3 (**Fig. 2E, H**). Interestingly, the microglia increased fraction was higher in the SN compared to other midbrain regions (**Fig. 2E upper panel; fig. S2 and table S7**). Moreover, image analysis of the microglia cellular shape (**fig. S2D**) revealed an IPD-related decrease in microglial branching exclusively in the SN, indicating cellular activation^25^ (**Fig. 2E lower panel** and **table S8**). We also confirmed the reduced fraction of oligodendrocytes in IPD (**fig. S3, table S6**) by quantitative immunofluorescence analysis targeting the PLP1 protein (**Fig. S4**). We detected a significant reduction of the PLP1-positive area and PLP1-intensity in the IPD3 midbrain sub-areas SN, and T/T compared to C3 (**fig. S4, table S7**). In contrast, the other midbrain cell types, OPCs, pericytes, ependymal, excitatory, inhibitory, and GABAergic cells, did not display significant differences associated with IPD (**fig. S3**). To our surprise, also, no loss in the number of DaNs was evident in the IPD samples, which is thought to be a critical factor in the pathogenesis of PD (**fig. S3**).

**Fig 2.**
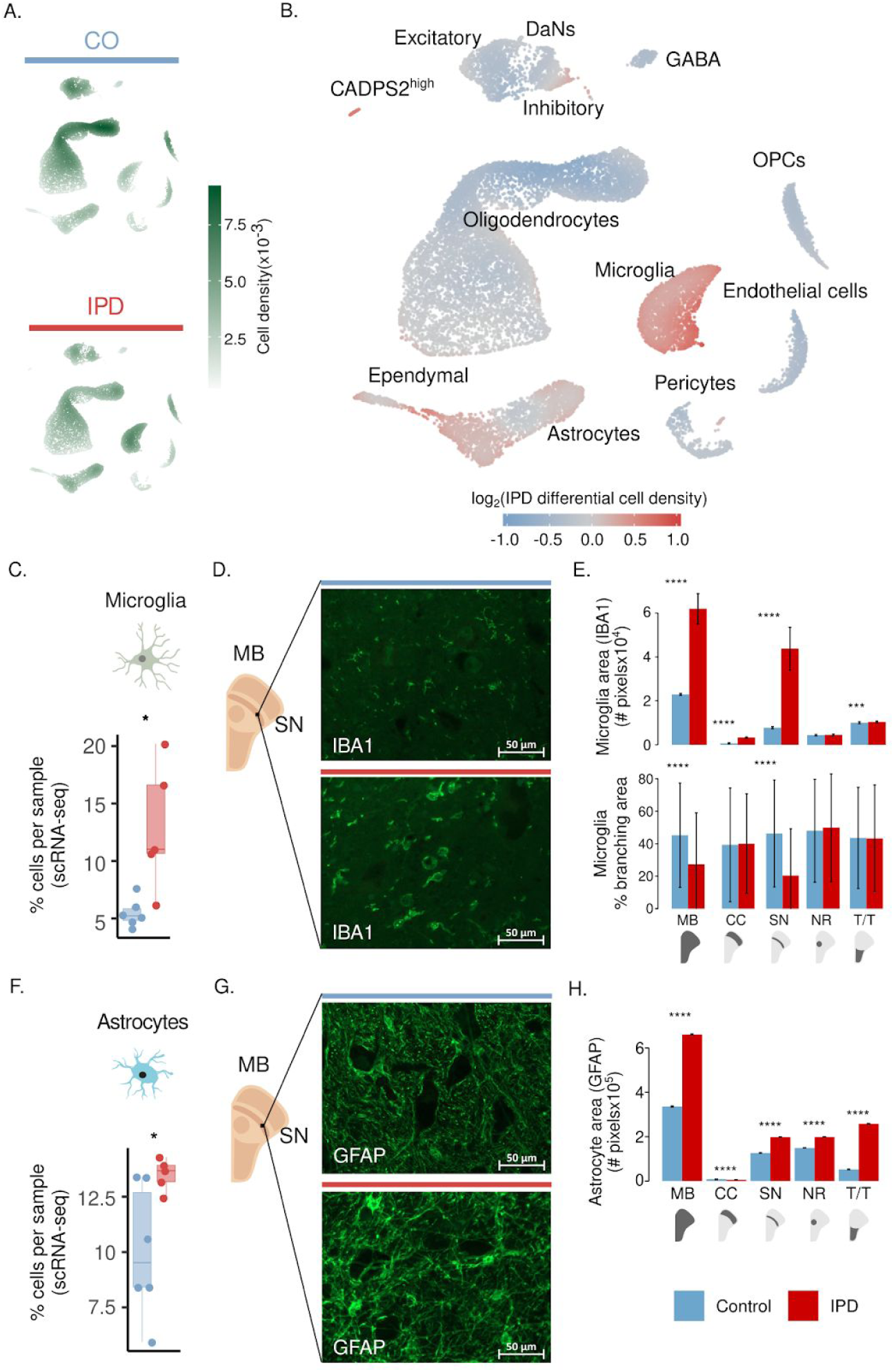
IPD midbrain is characterized by an increase in microglia and astrocytes. Differential cell-type composition in IPD patients compared to age-matched controls. (**A**) 2D cell density in the first UMAP embeddings of the human midbrain for IPD and control independently. (**B**) Differential 2D cell density in IPD midbrain. IPD midbrain has a larger population of microglia and astrocytes than control midbrain tissue. (**C**) Microglia cell proportion per sample. PD patients display a higher proportion of microglia cells (t-test *p*= 0.03). (**D**) IBA1 fluorescence in IPD3 and C3 in ventral midbrain sections. (**E**) Microglia area in the entire midbrain and individual regions. PD-associated increase of microglia is the most significant in the SN (t-test *p* < 0.0001). (**F**) Microglia IPD differential morphology. IPD-associated reduction of microglia branching indicates less ramified microglia which coincides with cell reactivity. (**G**) Astrocyte cell proportion per sample. An increase in the number of astrocytes was observed in IPD versus control midbrain sections (t-test *p* = 0.03). (**H**) GFAP fluorescence in C3 and IPD3 ventral midbrain sections. (**I**) Astrocyte area in the entire midbrain and individual regions. PD-associated increase of astrocytes is pronounced in each region (t-test *p* < 0.0001). MB, midbrain; SN, substantia nigra; NR, nucleus ruber; T/T, tectum/tegmentum; and CC, crus cerebri. PD, red bar; control, blue bar; scale bar = 50μm (* p < 0.1, ** p < 0.01, *** p < 0.001, **** p < 0.0001).

A closer look at the number of nuclei profiled indicated that DaNs only comprised 0.18% of the total cell count impeding a reliable comparison between IPD patients and controls (**Fig. 1F**). Therefore, we performed quantitative immunofluorescence imaging analysis of neuromelanin-positive areas in IPD3 and C3 and could confirm a 45.9% reduction in neuromelanin-positive nigral DaNs in IPD patients compared to controls (**fig. S5A; table S9)**. Moreover, in our study, 15 µm tissue sections, rather than tissue blocks, were used to isolate nuclei from control and IPD midbrain tissue. With a diameter of 10-20 µm, DaN nuclei are among the largest in the brain. Thus, when analysing nigral sections of IPD3 and C3 for TH-positive areas with and without nuclei, it became apparent that only a small subset of DaNs also contained (intact) nuclei (**fig. S5B, C**). This technical limitation likely contributed to the low number of DaNs detected by snRNA-seq. To validate DaN degeneration in the IPD tissue with an independent approach, we quantified the area of MAP2+/TH+/NM+ and MAP2+/TH-/NM+ neurons (**fig. S5D**). This analysis confirmed a significant decrease in DaNs and revealed an increase in the abundance of neuromelanin-containing neurons positive for MAP2 but negative for TH in IPD.

We also investigated how other clinical characteristics, in addition to condition (IPD), affect the midbrain cellular composition. For this, we modeled the percentage of each cell-type as a function of age, PMI, sex, and condition. We used beta-regression modeling to estimate the coefficients of these clinical features (**fig. S6**). Sex and condition (IPD) appeared to be the sample characteristics with the highest impact on the midbrain cellular composition. For instance, the most significant coefficients were the loss of DaNs and the gain of CADPS2^high^ cells associated with IPD (**fig. S6**). Male sex was negatively associated with the neuronal cells, inhibitory, excitatory, GABA and DaNs. Also, negatively related to the fraction of ependymal cells. Microglial abundance, in addition to the IPD condition, was positively associated with male sex. Together, these data suggest that PD pathophysiology might affect the midbrain cellular composition in a sex-dependent manner.

To assess if the increased fraction of microglia and astrocytes in IPD is associated with a specific molecular phenotype, we sub-clustered these cell-types to independently identify cellular subpopulations and reconstruct their activation trajectories. We identified seven microglia subpopulations characterized by the expression of a few marker genes (**Fig. 3A**). The three biggest subpopulations are defined by the high expression of P2RY12, GPNMB, and HSP90AA1 (**Fig. 3B**). Given that these three sub-populations conform a continuum in the UMAP projection and both GPNMB and HSP90AA1 genes are markers of microglia activation, we estimated a cell trajectory structure comprising these major sub-populations using the DDRTree method of monocle3 (**Fig. 3C**). We then organized cells along this trajectory (pseudotime), starting from the trajectory node that maximizes the distance to the GPNMB and HSP70AA1 trajectory branch tips (**Fig. 3C**). This microglia activation trajectory spans from P2RY12^high^ cells towards two activation branches, one containing GPNMB^high^ cells and another with cells highly expressing HSP90AA1 or IL1B (**Fig. 3C**). IPD cells differentially distribute along the microglia activation trajectory and IPD microglia tend to be in an activated state compared to controls (**Fig. 3D**). While P2RY12 is highly abundant in resting microglia^26^, GPNMB^27^, HSP90^28^ and IL-1β^29^ are involved in the inflammatory response and have previously been linked to neurodegeneration^28,30,31^, supporting the notion that the IPD-specific upregulation of *GPNMB* and *HSP90AA1* are makers of microglial activation. To further characterize the molecular phenotype of these two activated microglia states, we identified genes whose expression was associated with the activation trajectory and functionally enriched them to gene-ontology molecular functions (**Fig. 3D, E**).

**Fig 3.**
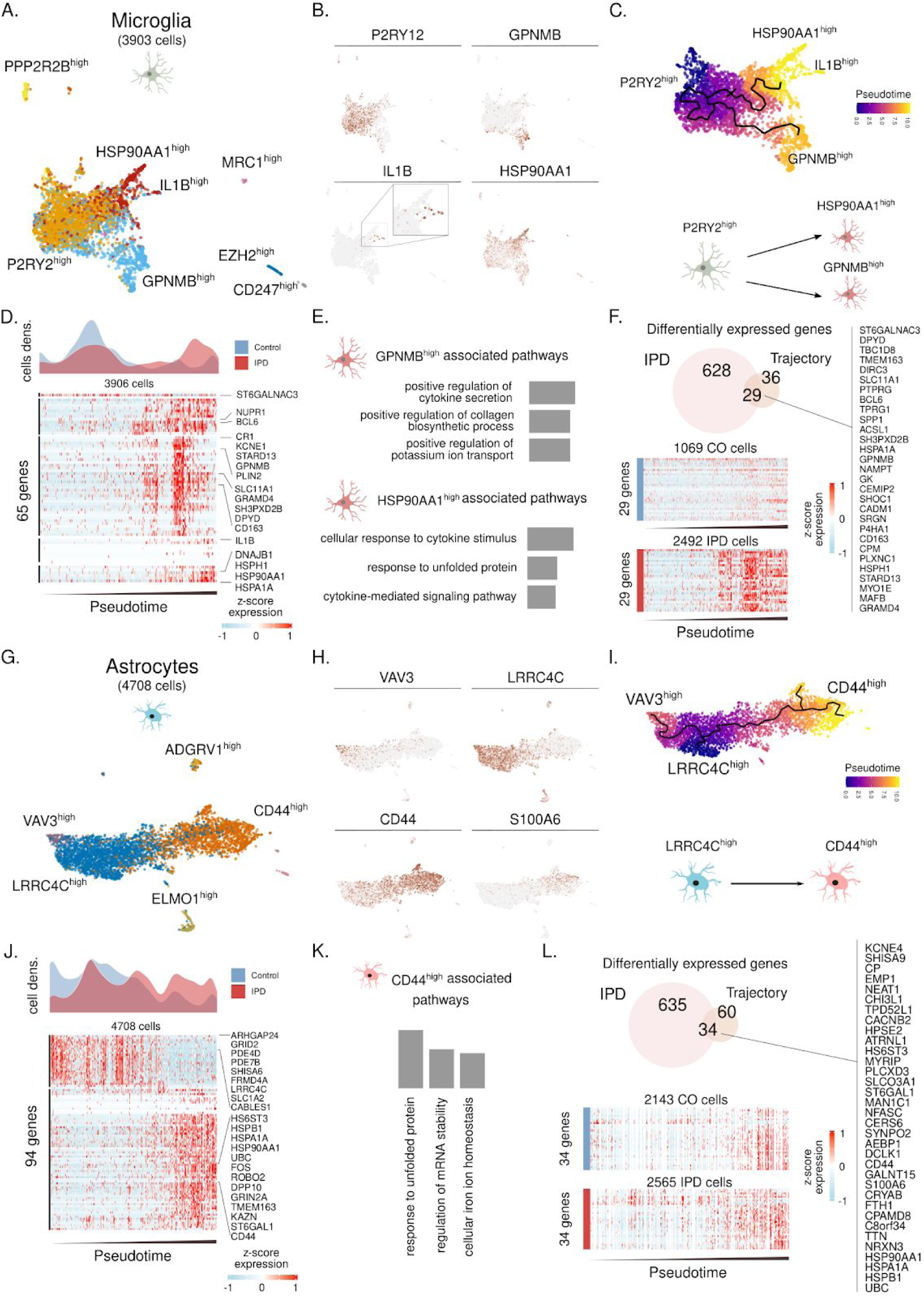
Trajectory reconstruction reveals microglia and astrocyte differential activation in IPD. (**A**) Microglia subpopulations labeled with a representative marker gene. (**B**) Expression of *P2RY12, GPNMB, IL1B* and *HSP90AA1* along the nearly four thousand microglia cells. These genes are characteristic of the major three microglia subpopulations. (**C**). Trajectory reconstruction and pseudotime representation based on the P2RY1^high^, GPNMB^high^, and HSP90AA1B^high^ subpopulations. This reveals a two-branches activation trajectories. (**D**) IPD and control differential cell-density distribution along pseudotime. Also, the expression of 65 genes whose expression is associated with the microglia activation trajectory. Z-score normalized expression is presented for each gene over nearly four thousands microglia cells organized by their pseudotime. (**E**) Gene ontology (GO) molecular function enrichment of the genes associated with the *GPNMB* and *HSP90AA1* activation trajectories. (**F**) 29 IPD-differentially expressed genes intersect with the differentially expressed genes along the microglia activation trajectory. (**G**) Astroglial subpopulations named based on characteristic marker genes. (**H**) *VAV3, LRRC4C, CD44* and *S100A6* expression across the ∼4700 astrocytes. (**I**) Inferred cell trajectory and pseudotime for the major astrocyte subpopulations, VAV3^high^, LRRC4C^high^, and CD44^high^ cells. (**J**) IPD and control differential cell-density distribution over pseudotime and the expression of the 94 genes highly associated with the astrogliosis trajectory in the ∼4700 astrocytes organized by pseudotime. (**K**) GO molecular function pathway enrichment of the upregulated genes in the CD44^high^ activated branch. (**L**) The 34 intersected genes between the upregulated genes in IPD and across the astrocyte activation trajectory.

This analysis revealed that these subpopulations are involved in the secretion of cytokines and the stress response to unfolded proteins (**Fig. 3E**). Next, we identified the genes whose expression was differentially upregulated in IPD microglia across the activation trajectory. We intersected the upregulated genes and the activation-trajectory associated genes in microglia and identified 29 genes linked with the IPD differential activation of the midbrain microglia (**Fig. 3F**), several of which have previously been associated with IPD^32–34^.

Following this approach we also characterized the astrocytes subpopulations, reconstructed the astrocyte activation trajectory and identified gene signature associated with IPD differential activation (**Fig. 3G-L**). We identified five astrocytes subpopulations characterized by high expression of VAV3, LRRC4C, ELMO1, ADGRV1, and CD44 (**Fig. 3G-H**). We recovered the astrocyte activation trajectory based on the main cell types comprising VAV3^high^, LRRC4C^high^, and CD44/S100A6^high^ subpopulations (**Fig. 3I**). Given that CD44 expression implicates reactive astrogliosis^35^ we ordered cells on the activation trajectory by setting the root in the trajectory graph-node that maximizes the distance from the CD44^high^ branch end. These results implied an astrocyte activation transition from LRRC4C^high^ to high CD44^high^ subpopulations (**Fig. 3I**). Indeed, IPD astrocytes were highly enriched at the end of the astrogliosis trajectory compared to control astrocytes (**Fig. 3J**). We further characterized the molecular phenotype of the CD44^high^ astrocyte activated stated by enriching GO molecular functions to the highly upregulated genes across the astrocytes activation trajectory (**Fig. 3J-K**). The CD44^high^ subpopulation was related to the unfolded protein response (UPR) pathway, which has recently been linked to a specific astrocyte reactivity state that is detrimental to the survival of neurons^36^ **(Fig. 3K**). Next, we calculated IPD-differentially upregulated genes, which were also highly expressed towards the end of the astrogliosis trajectory (**Fig, 3L**) and identified 34 genes associated with IPD differential astrogliosis (**Fig. 3L**). These genes include several heat-shock proteins that have previously been shown to co-localize with alpha-synuclein deposits in the human brain^37^.

Finally, with this cell-type-specific approach, we investigated the oligodendrocyte diversity and reconstructed its differentiation trajectory (**Fig. S7**). We identified five subpopulations characterized by the expression of *ATP6V02, OPALIN, TRPM3, ST6GAL1*, and *RBFOX1* (**fig. S7A-B**). The inferred trajectory based on subpopulations recovers differentiation trajectory spanning from FRY/OPALIN^high^ cells towards RBFOX1/S100B^high^ cells (**fig. S7B-C**). *OPALIN* (also named as Tmem) is a marker of myelinating oligodendrocytes^38^, while S100B has been associated with glial stress response in PD postmortem midbrain^39^. When comparing IPD oligodendrocyte density across this trajectory, we found a reduced fraction of myelinating OPALIN^high^ cells compared to controls (**Fig. S7D**). An overlay of the IPD differentially expressed genes and such defining the oligodendrocyte trajectory identified 216 and 330 down-regulated and upregulated genes in the IPD and across the trajectory. Downregulated genes are associated with neuronal maintaining pathways, while upregulated genes are related to the response to unfolded protein pathways (**fig. S7E-F**).

We then focused on deciphering the identity of the CADPS2^high^ cells, which were almost exclusively detected in the IPD patients (**Fig. 4A**). This cell population pseudo-bulk-transcriptome clusters together with the neuronal cell types, excitatory, inhibitory, GABAergic, and DaNs (**fig. S1I**). In accordance with this global transcriptome similarity, CADPS2^high^ cells positively express the neuronal marker genes MAP2 and SCN2A (**Fig. 4B**). Performing immunofluorescence labeling with antibodies for CADPS2, MAP2, and TH, we confirmed the existence of a neuronal cell population with an IPD-specific increase of CADPS2 expression in the SN (**Fig. 4C, E**). Primary, SN is the midbrain region where we detected a CADPS2 intensity higher in IPD patients than control subjects (**Fig. 4C**). Therefore, we then characterized the CADPS2 intensity in the SN neurons. We found that CADPS2 intensity was higher in IPD than controls in the TH-negative neurons harboring neuromelanin deposits (MAP2+/TH-/NM+) (**Fig. 4D-E**). Given that the TH positive neurons (MAP2+/TH+/NM+) do not display CADPS2 intensity differences (**Fig. 4D**), we concluded the CADPS2^high^ cells represent nonfunctional dopamine neurons. In accordance with their snRNA-seq profiles, cells with high CADPS2 expression were positive for MAP2 but had low TH levels (**Fig. 4B, E**). Besides, CADPS2^high^ cells showed high *TIAM1* expression (**Fig. 4B**), a key regulator of neuronal differentiation of dopaminergic precursor cells^40^. In agreement with this finding, cell-cycle labeling analysis indicates that while the majority of excitatory, inhibitory, GABAergic, and DaNs neurons were in a resting state (G0/G1), a considerable proportion of CADPS2^high^ neurons had entered the cell cycle (G2/M/S) (**Fig. 4F**). We then co-embedded the DaN and CADPS2^high^ cell populations with the developmental trajectory of the human dopamine neurons (6-11 weeks-old). The DaNs from our study predominantly clustered with differentiated DaNs (**Fig. 4G, H**) while the CADPS2^high^ neurons were transcriptionally more similar to embryonic neuroblasts (**Fig. 4G, H**). These results might indicate a cell cycle re-entry and partial re-activation of the developmental machinery in CADPS2^high^ neurons.^41^

**Fig 4.**
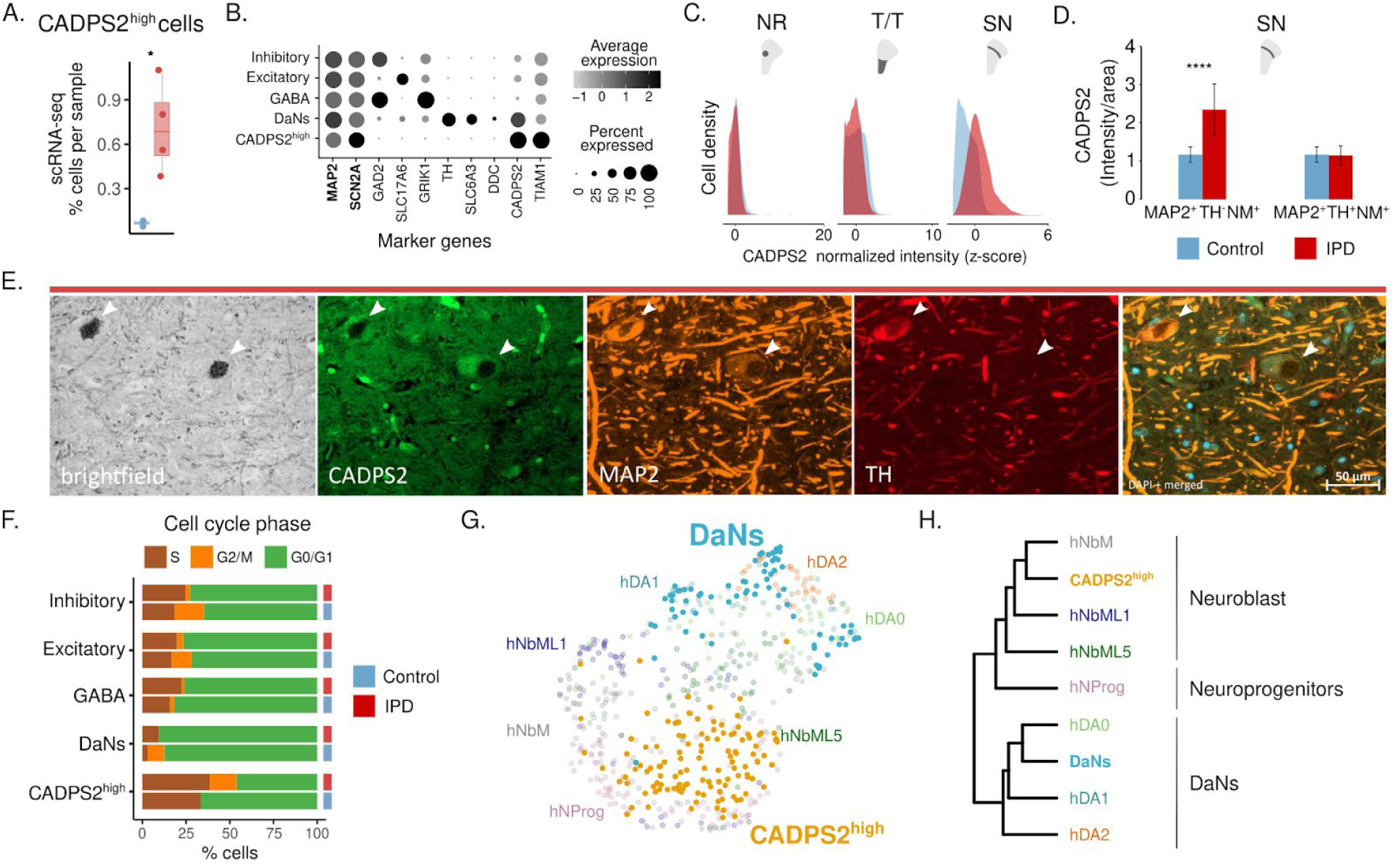
CADPS2^high^ cells are neurons that have lost their dopaminergic potential and try to advance through the cell cycle. (**A**) CADPS2^high^ cell proportion per sample, (t-test *p* = 0.02). (**B**) CADPS2^high^ cells are neurons. They express *MAP2, SCN2A* and *TIAM1* but have low levels of *TH*. (**C**) When comparing different subregions of the midbrain using immunofluorescence analysis, only in the SN, CADPS2 protein was more abundant in IPD than in control tissue. (**D**) Average CADPS2 fluorescence intensity in MAP2^+^/TH^+^/NM^+^ and MAP2^+^/TH^-^/NM^+^ neurons in C3 and IPD3 ventral midbrain sections, indicating enhanced CADPS2 levels in IPD DaNs with low *TH* levels. (**E**) Representative image of MAP2^+^/TH^-^/NM^+^ and MAP2^+^/TH^+^/NM^+^ cells in the SN of the IPD3 patient. MB, midbrain; SN, substantia nigra; NR, nucleus ruber; T/T, tectum/tegmentum; and CC, crus cerebri. IPD, red bar; control, blue bar; scale bar = 50μm (* p < 0.1, ** p < 0.01, *** p < 0.001, **** p < 0.0001). (**F**) Cell cycle phase assignment proportions. (**G**) UMAP co-embedding of the snRNA profiles of DaNs and CADPS2^high^ cells from the current study with the human developmental trajectory of dopamine neurons^41^. (**H**) While the DaNs from postmortem IPD and control midbrain tissue cluster together with the embryonic dopamine neurons (hDA0-0/1/2), CADPS2^high^ cells cluster with embryonic neuroblasts (hNbML/1/5) and neural progenitors (hNprog).

## Discussion

In this study, we describe a single-cell atlas of the human midbrain from IPD patients and age-matched controls. Applying snRNA-seq to postmortem midbrain tissue, we profiled the transcriptomes of more than 41,000 single-nuclei with the aim to identify cell type- and disease-specific molecular signatures associated with IPD.

In the current dataset glia made up ∼80% of all sequenced cells, enabling an in-depth analysis of their contribution to the pathogenesis of IPD. We identified a disease-specific upregulation of microglia, which mediate the innate immune defense in the brain. During microgliosis, microglia amplify, secrete cytokines and undergo morphological changes^42^. Suggestive of an activated state, we detected less ramified microglia in IPD postmortem tissue using a quantitative immunofluorescence approach. Moreover, we identified a significant PD risk variant enrichment in microglia showing the strongest association with the PD gene *LRRK2*. The kinase LRRK2 is most abundant in immune cells and may contribute to inflammasome formation via the phosphorylation of Rab GTPases^43^. In addition, by inferring the activation trajectories of the microglial subpopulations, we observed a transition of cells from resting into an activated state. In agreement with this finding, pathway analyses highlighted cytokine signalling and, likely upstream of this, an induction of the UPR pathway. We also found chaperones and heat-shock proteins overexpressed along the disease trajectory, which when they are released from the cell can act as damage-associated molecular patterns (DAMPs) that trigger an immune reaction^44^.

Astrocytes can equally act as immune effector cells in the brain by releasing proinflammatory cytokines^45^, likely explaining why we observed a PD risk variant enrichment in these cells. Furthermore, when modelling astroglial activation trajectories, we detected reactive astrogliosis specifically in patient cells^35^. As for microglia, pathway analysis along the trajectory identified the UPR pathway, which has recently been described to influence the astrocytic secretome.^36^ Neurotrophic factors released from reactive astrocytes were shown to accelerate neuronal demise^36^ - a disease mechanisms that has not gained much attention in PD research so far.

Our snRNA-seq data also showed a trend towards decreased oligodendrocyte numbers in IPD midbrain - a finding that we could validate by immunohistochemistry. In the white matter, oligodendrocytes generate myelin sheets, which provide insulation of axons and ensure saltatory conduction^46^. However, since PD has long been considered a “gray matter” disease, oligodendrocytes only recently gained attention in the field. Combining the latest genome-wide association study (GWAS)^47^ with snRNA-seq results from IPD and control midbrain sections, we did not observe a significant PD risk association for oligodendrocytes.

However, trajectory inference analysis revealed a transition from high *OPALIN* to high *S100B* expression subpopulations. While S100B was shown to control the maturation process of oligodendrocytes^48^, the protein has also been linked to neurodegeneration. *S100B* overexpression in response to cytokine injections mediates dystrophic neurite formation in an Alzheimer rat model^49^. Accordingly, the oligodendrocyte-specific upregulation of *S100B* observed in the IPD midbrains may be the result of enhanced cytokine release from microglia. Taken together, these results further implicate glial cells in the propagation of neuroinflammatory and neurodegenerative processes in IPD.

When focusing on DaNs in our snRNA-seq study, we did not observe a significant loss in IPD tissue. However, a meaningful comparison in IPD patients and controls was likely hampered by the low abundance of DaNs in general - they only constituted 0.18% of the total cell count. By contrast, automated image analysis of immunofluorescence labelled IPD and control midbrain sections indicated a significant loss of neuromelanin containing DaNs. This result was in line with the neuropathological reports, which described severe DaN degeneration in all IPD patient samples (cp. **table S1**). Thus, technical limitations may have caused the underrepresentation of DaNs in the transcriptomic data: first, we used midbrain slices of a thickness of ∼15µm. By contrast, with an average diameter of 10-20 µM, DaN nuclei are rather large. Hence, a considerable proportion of nuclei may not have remained intact during the sectioning process - a prerequisite for high-quality snRNA-seq results. Second, when we previously applied scRNA-seq to midbrain organoids, the detected number of TH-positive neurons was well below the actual DaN count^50^. One possible explanation for this discrepancy could be that the majority of *TH* mRNA transcripts lack or have a very short poly(a) tail and are therefore not recognized by snRNA-seq^51^. Despite these constraints, our cell-type specific analysis for PD variants in midbrain tissue revealed an enrichment in DaNs.

Finally, we discovered a new disease-specific cell state that is defined by translational similarity to midbrain DaNs but with low *TH* levels and high *CADPS2* expression. Immunofluorescence analysis revealed that CADPS2^high^ neurons are localized to the SN and that they harbour neuromelanin deposits. CADPS2 (Calcium-dependent activator protein for secretion 2) functions in the uptake and storage of vesicular monoamines^52,53^ such as dopamine and has previously been linked to genetic PD^54^. In the presence of the LRRK2 mutation G2019S, *CADPS2* expression was found to be enhanced^54^. However, in our patient samples LRRK2 mutations were excluded. In addition to high levels of *CADPS2*, the IPD-specific neuronal population was characterized by elevated *TIAM1* expression. TIAM1 (T-cell lymphoma invasion and metastasis 1) regulates midbrain DaN differentiation via the Wnt/Dvl/Rac1 pathway^40^. Wnt signalling is crucial in the development of DaNs and has become a major target for regenerative treatment approaches in PD^55^. Accordingly, increased *TIAM1* expression in CADPS2^high^ neurons may suggest that these neurons revert to an earlier developmental stage. This is in line with our observation that, unlike DaNs, a large proportion of CADPS2^high^ neurons was found in a non-resting state. An incomplete cell-cycle re-entry of DaNs has previously been reported in postmortem brain samples from IPD patients^41^. Further, this study showed that the inability to re-start a developmental program mediates DaN demise^41^. Here, we may have observed a similar phenomenon: DaNs that are highly metabolically active accumulate neuromelanin in an accelerated fashion until a certain threshold is reached. Beyond this tipping point, the neuromelanin deposits become detrimental^56^, resulting in a partial activation of the developmental machinery and, subsequently, the death of these DaNs. Supporting this hypothesis, CADPS2^high^ neurons clustered with neural progenitor cells and neuroblasts when we co-embedded our single-cell results onto scRNA-seq data from human embryos. Thus, we speculate that the TIAM1/CADPS2^high^ population of cells represents DaNs that are in the process of dying.

In summary, our study revealed several novel aspects of IPD pathology. Using snRNA-seq in postmortem midbrain tissue from patients and matched controls, we identified a disease-specific upregulation of microglia and astrocytes as well as a loss of oligodendrocytes. In addition, we discovered a novel neuronal cell state that was almost exclusively identified in IPD midbrain tissue. These cells were characterized by low *TH* levels but high expression of *CADPS2* and *TIAM1*. Based on our scRNA-seq and quantitative imaging data, we conclude that these neurons lost their dopaminergic identity and unsuccessfully attempted to re-enter the cell cycle. Our findings strengthen the role of neurodevelopmental and neuroinflammatory mechanisms in the pathogenesis of PD opening new avenues for novel therapeutic strategies.

## Materials and Methods

### Human brain tissue cryosectioning

Frozen human postmortem midbrain tissue sections and associated clinical and neuropathological data were supplied by the Parkinson’s UK Brain Bank and the Newcastle Brain Tissue Resource. According to the neuropathological procedure, after removing the brainstem and cerebellum, the brain hemispheres were divided down the midline with the hemi-midbrain associated with each hemisphere. The left hemi-midbrain was removed with a transverse section by taking a line from just behind the mammillary body through the superior colliculus. This midbrain block was then snap-frozen at −120°C and cryosectioned at ∼15 μm thickness in the transverse plane. The resulting sections were stored at −80°C.

Patients and controls gave written informed consent with the brain banks which, together with the ethics review panel of the University of Luxembourg, approved the study.

### Sample preparation for nuclei isolation

Six to eight sections were combined from one individual for nuclei isolation. Nuclei were isolated by adapting the published 10X Genomics^®^ protocol for ‘Isolation of Nuclei for Single Cell RNA Sequencing’ In brief, the tissue was lysed in a chilled lysis buffer (10 mM Tris-HCl, 10mM NaCl, 3mM MgCl_2_, 0,1% NonidetTM P40). Then, the suspension was filtered and nuclei were pelleted by centrifugation. Nuclei pellets were then washed in ‘nuclei wash and resuspension buffer’ (1xPBS, 1% BSA, 0.2U/μl RNase inhibitor), filtered and pelleted again. Nuclei pellets were suspended in DAPI solution (1.5 μM DAPI in 1xPBS) and incubated for 5 minutes prior to sorting. After dissociation, single DAPI-positive nuclei were FACS-sorted using a FACSDiva Cell Sorter (BD Biosciences).

### Library preparation and sequencing

Sorted nuclei were processed using the Chromium Next GEM Single Cell 3’ Kit v3.1 to generate the cDNA libraries. The quality of cDNA was assessed using the Agilent 2100 Bioanalyzer System. Sequencing was performed on Illumina NovaSeq 6000-S2.

### Transcript quantification and filtering

FASTQ files were generated from the raw base call (BCL) outputs with the Cell Ranger (10X Genomics) *mkfastq* pipeline v.3.0. From this, we obtained a gene-barcode UMI *count* matrix per sample using the Cell Ranger (10X Genomics) count pipeline v.3.0 using default parameters. The Cell Ranger count pipeline only considers exon-mapping reads during UMI-counting. Also, single-nuclei sequencing readouts are enriched in intronic regions. To account for this, we used the Cell Ranger recommended variation of the human reference transcriptome (hg38), where introns are annotated as exons. We retained barcodes with more than 1500 UMIs and 1000 genes, as well as less than 10% of mitochondrial-encoded (mtDNA) and 10% of ribosomal gene counts. We kept genes detected in more than three barcodes. Also, we filtered out ribosomal and mtDNA-encoded genes. We then used Scrubblet^57^ to identify potential multiplet barcodes. We kept barcodes with an estimated duplet score smaller than 0.15 for downstream analysis.

### Normalization, sample integration, and cell clustering

To identify the major cell types comprising the human midbrain, we combined the samples in a single embedding following the Seurat v3^58^ integration workflow. First, each sample was normalized using the SCTransform approach^59^. Cell-cycle phase assignment was performed based on this normalized expression matrix. We used the Seurat *CellCycleScoring* function and the Seurat v3 reference genes for the S and G2/M cell-cycle phases. To determine the between-sample cell-anchors, we used the *FindIntegrationAnchors* Seurat function with the top 4000 consistently, most variable genes in all samples, which were identified with the *SelectIntegrationFeatures* function. We then used the *IntegrateData* Seurat function to obtain a combined and centered expression matrix. Principal component (PC) analysis was done on this centered expression matrix. The top 25 PCs were used to build a Shared Nearest Neighbor (SNN) cell graph, which was then clustered using the Louvain algorithm (resolution = 1.5) implemented in the Seurat *FindClusters* function. The top 25PCs were embedded into two dimensions using the Uniform Manifold Approximation and Projection (UMAP) algorithm^60^. We identified marker genes for each cluster by using the ROC method of the Seurat *FindAllMarkers* function. The top marker genes were used to assign cell-type annotations manually for each cell cluster. We compared the cell types by correlating their pseudo bulk profiles. The resulting gene-cell-type matrix was normalized (Transcript Per Million) and log2 transformed. The Pearson correlation estimates among the normalized cell-type profiles were used as the input distance matrix for hierarchical clustering.

### Differential cell-type composition

We estimated the differential cell type composition by comparing the UMAP embeddings and the cell type proportions between IPD and control samples. We considered the two-dimensional kernel cell density of the IPD and control cells independently on the first two UMAP components using the *kde2d* function (bins = 100) implemented in the *MASS* R package^61^. The IPD log2 differential UMAP density was calculated. Also, for each cell type, we compared the proportion of cells per sample between the IPD patients and control individuals. We assessed this difference with the Student’s t-test implemented in the *t*.*test* function of the R stats package^62^. Furthermore, we used the beta-regression model to estimate the contribution of the sample clinical features (e.g., condition, postmortem interval [PMI], age, sex) on the cell proportion variation. We modeled the cell type proportion using the betareg R package^63^.

### Sub-clustering, trajectory reconstruction, and differential gene expression in microglia, astrocytes and oligodendrocytes

We subset cell-type-specific UMI raw counts. To identify the cell type subpopulations, we integrated cells from different samples following the Seurat3 Reciprocal Principal Component Analysis based protocol considering the top 1000 highly variable genes for astrocytes and oligodendrocytes or the top 500 highly variable genes. Then we used the unsupervised and network-based Louvain clustering approach based on the top 25 principal components of the integrated datasets. Marker genes were defined as described before. We reconstructed the cellular activation trajectories following the monocle3 approach. Briefly, cells from different samples were integrated and factor size normalized. The sample effect was removed using the Mutual Nearest Neighbor method^64^. Then the highly variable genes defined before were embedded in the first 25 principal components used for dimensionality reduction and trajectory inference using the *learn_graph* function implemented in the monocle v3 R package^8^. Pseudotime ordering was done in a supervised manner by rooting the trajectory in the graph node that maximizes the distance to the known activated cell subpopulation. We identified cell-type-specific perturbed genes in IPD by fitting negative binomial distributions to each gene using the *fit_models* function implemented in the monocle v3 R package^8^. IPD differential expression coefficient with *q* < 0.05 were considered as differentially expressed genes. Highly variable genes associated with the cell trajectories were identified by using the spatial correlation analysis Moran’s I approach implemented in the *graph_test* function of the monocle v3 R package^8^. Functional enrichment analysis of the differentially perturbed genes was done using Enrichr^65^.

### Human embryonic and adult dopamine neurons co-embedding

We merged the raw UMI count matrix comprising the human developmental trajectory of dopamine neurons available in GSE76381 GEO accession and the UMI count matrix containing the adult dopamine and CADPS2^high^ neurons reported in this study in a single dataset. To remove the study effect, we followed the Seurat3 Reciprocal Principal Component Analysis based Seurat3 integration protocol using the embryonic cells as reference and considering the top 4000 highly variable genes among the embryonic cells. Based on the integrated top 25 Principal Components, we embedded these cells into two UMAP dimensions. Cluster analysis of these cell-types was performed using the *BuildClusterTree* implemented in Seurat3.

### Automated image analysis

Immunofluorescence images of human postmortem midbrain sections were acquired with Carl Zeiss Axio Observer Inverted Microscope Z1 and analyzed in Matlab (Version 2019B, Mathworks). Automated in-house developed image analysis algorithms segment the fluorescent cell areas (neurons, astrocytes, microglia, oligodendrocyte) extracting features such as area, perimeter and mean fluorescence intensity. The segmentation of all neurons was computed by convolving the raw MAP2 channel with a Gaussian filter. By setting a pixel threshold, all high intensity neurite areas were detected and subtracted from the image. The MAP2 mask was generated by setting the pixel threshold, followed by bwareaopen to remove small connected components. The detected areas were further segmented with watershed to dissociate small objects from the cell areas, which are removed by another bwareaopen filter in order to generate the final MAP2 mask. The segmentation of dopaminergic neurons was computed by convolving the raw TH channel with a Gaussian filter. By setting a pixel threshold, TH-positive cell areas were detected followed by bwareaopen to remove small connected components in order to generate a TH area mask. The neuromelanin mask was computed by identifying areas below the selected pixel threshold and removing the small connected components with bwareaopen. In addition, we identified the MAP2+/TH+/NM+ and MAP2+/TH-/NM+ area masks by subtracting and adding the respective individual mask areas. The protein level of CADPS2 was identified by overlaying a respective mask (MAP2+, MAP2+/TH+/NM+ and MAP2+/TH-/NM+ mask) over the raw CADPS2 channel and detecting the mean intensity of the pixels in the mask area. Each measured intensity was then normalized to the area size. The segmentation of astrocytes and microglia was computed by selecting a pixel threshold, followed by an imfill filter in order to generate the cell area masks for GFAP or IBA1, respectively. Further, the skeleton of the IBA1 mask was generated with a thinning function to identify the branching of the mask. Because of the massive population of oligodendrocytes, we generated the mask by selecting a pixel threshold to identify all the PLP1 positive areas without segmentation. The results were visualized with ggplot2 in R 4.0.0.

### Genotyping of PD cases using NeuroChip

DNA samples from all PD cases underwent genotyping at the Institute of Human Genetics at the Helmholtz Zentrum München using the Illumina (San Diego, CA) NeuroChip^66^. Standard genotype data quality control (QC) steps were carried out^67^. SNP imputation was carried out on our NeuroChip data using the Michigan Imputation Server^68^ to produce a final list of common (minor allele frequency ≥ 1%) variants for further analyses. Imputed SNP positions were based on Genome Reference Consortium Human 37/human genome version 19 (GRCh37/hg19). All cases were screened for disease-associated variants in known major PD genes (*SNCA, LRRK2, DJ-1, PRKN, GBA, PINK1, ATP13A2, VPS35, MAPT, DCTN1, DNAJC6, SYNJ1, VPS13C* and *MAPT*) covered by the NeuroChip.

### Cell type association with genetic risk of PD

Association analysis of cell type-specific expressed genes with genetic risk of PD was performed using Multi-marker Analysis of GenoMic Annotation (MAGMA) v1.07, in order to identify disease-relevant cell types in the midbrain. MAGMA is a gene set enrichment analysis method, which tests the joint association of all SNPs in a gene with the phenotype, while accounting for LD structure between SNPs^69^. Competitive gene set analysis was performed on SNP p-values from the GWAS summary statistics of Nalls et al. (excluding 23andMe)^47^ and the publicly available European subset of 1000 Genomes Phase 3 was used as a reference panel to estimate LD between SNPs. SNPs were mapped to genes using NCBI GRCh37 build (annotation release 105). Gene boundaries were defined as the transcribed region of each gene. An extended window of 35 kb upstream and 10 kb downstream of each gene was added to the gene boundaries^70^. ZSTAT is the Z-score (mean zero and unit SD) for the gene, based on its p-value. It was used to determine the gene association in the gene level analysis. If a gene is not differentially expressed, Z-scores will have approximately zero mean and unit variance. Gene sets used in MAGMA were filtered for: FDR-corrected p-values < 0.05, percentage of cells of the cluster where the expression was detected > 0.5, and logFC > 0.25.

### Machine learning cross-validation of cell type annotation

To quantitatively assess the cell-cluster definition and annotation, we implemented a stratified cross-validation machine learning approach. Briefly, we removed the sample effects on the combined UMI count dataset using Harmony^71^. For normalization, we used the loess transform^72^ to fit a smooth curve between mean and variance using the log transformed data. We then scaled the data with the fitted mean and standard deviation. The identified marker genes (**Table S3**) were selected as the features of the model. We considered the median cell number of each cell-type to subsample the dataset as we had few cell-types (DaNs and CADPS2^high^) underrepresented. To ensure equal label composition in the training and the test sets, we split the data using scikit-learn^73^ *StratifiedKFold* with 70% of the data as training and 30% as test dataset and a 5-fold cross validation. We did a dimensional reduction with *truncatedSVD* to 30 components. These 30 components were classified based on scikit-multilearn’s ensemble classification^74^ which uses louvain-based clustering^75,76^ and a random forest classification to account for the clustered and sparse nature of the scRNAseq data. The predicted cell types were then compared to the manually curated cell label assignments using a confusion matrix.

## Data Availability

Raw data for the 11 samples presented in this study are available in the Gene Expression Omnibus (GEO) with accession number GSE157783. Single-cell human embryonic midbrain dataset was available in the GSE76381 GEO accession.

https://www.ncbi.nlm.nih.gov/geo/query/acc.cgi?acc=GSE157783

## General

We express our gratitude to the tissue donors and their families for their generous participation.

## Author contributions

S.Sm., C.A.P-M., A.G. and M.S. designed the study. Experiments were performed by S.Sm., C.D., S.Sa., J.J. and J.H.; sequencing was performed by B.T.; gene expression data was analysed by C.A.P-M and S. B.. Imaging data analysis was performed by S.Sm., S.P., J.J. and P.A.; tissue collection and neuropathological analysis was carried out by C.M.M.; IPD genetic profiling was performed by S.P., Z.L. and P.M. PD genetic variant enrichment was accomplished by Z. L., C.A.P-M., and P. M.. S.Sm., C.A.P-M., J.C.S., P.M., A.G. and M.S. wrote the manuscript; all authors contributed to the critical revision of the manuscript.

## Supplementary figures

**Fig S1.**
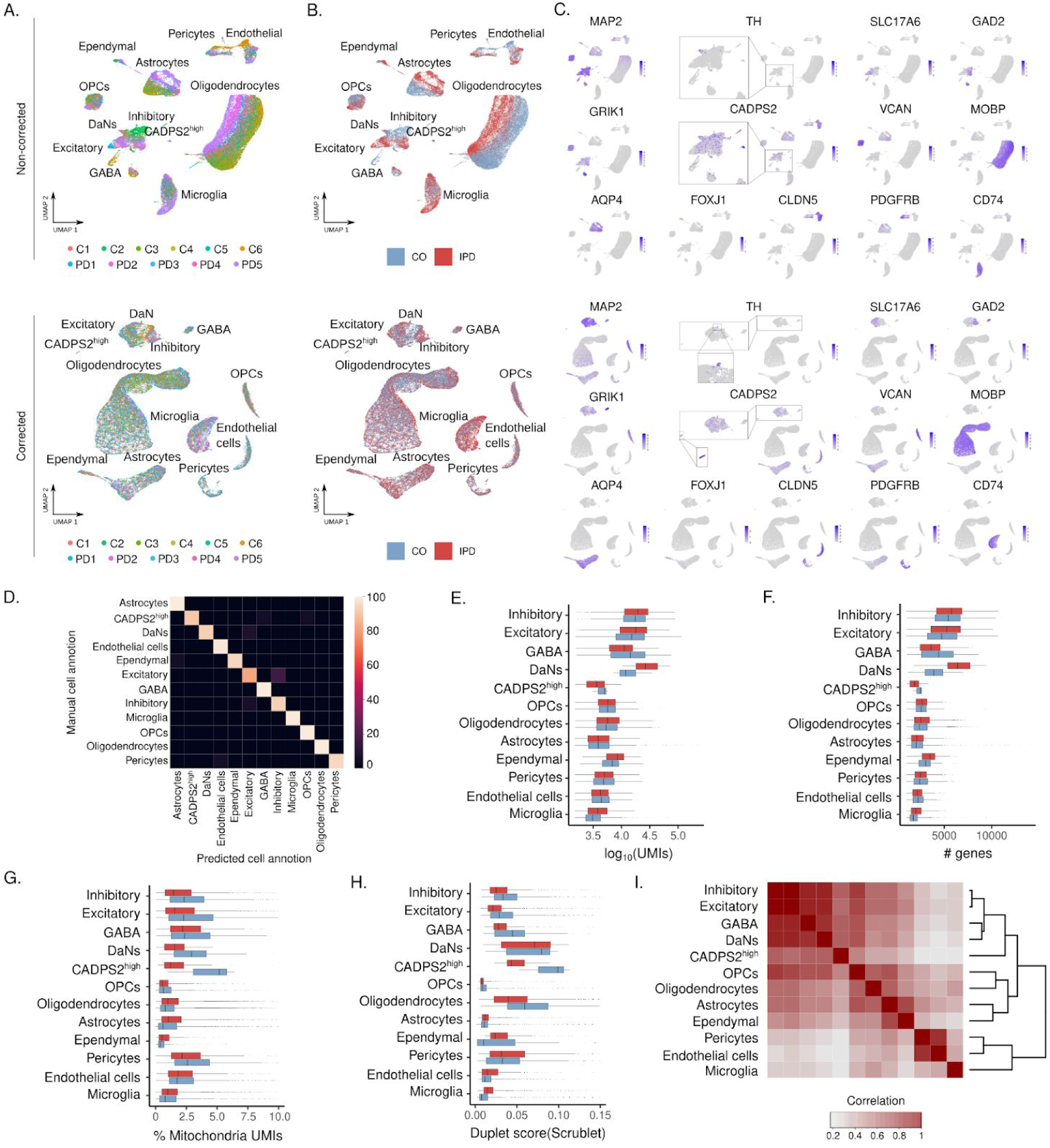
Midbrain cell UMAP embedding and cell-type-specific snRNA-seq quality control metrics and similarity. (A-B) Midbrain single-cell atlas UMAP embedding colored by sample and condition. Top panels display UMAP embeddings based on the top 25 non-corrected principal components. Bottom panels show the UMAP embeddings based on the top 25 principal components after removing the inter-individual variability using the Seurat3 Canonical Correlation Analysis based integration protocol. (C) Expression distribution of cell-type marker genes on the ∼41.000 midbrain cells. (D) Confusion matrix results of the machine learning cross-validation approach to validate the cell type definition. (E) UMI count distribution. (F) Number of detected genes. (G) Percentage of mtDNA-encoded transcripts per cell. (H) Scrublet duplet score. (I) Cell-type pseudo-bulk correlation matrix. CADPS2^high^ pseudo-bulk transcriptome profile clusters together with the neuronal cells, inhibitory, excitatory, GABA and DaNs.

**Fig S2.**
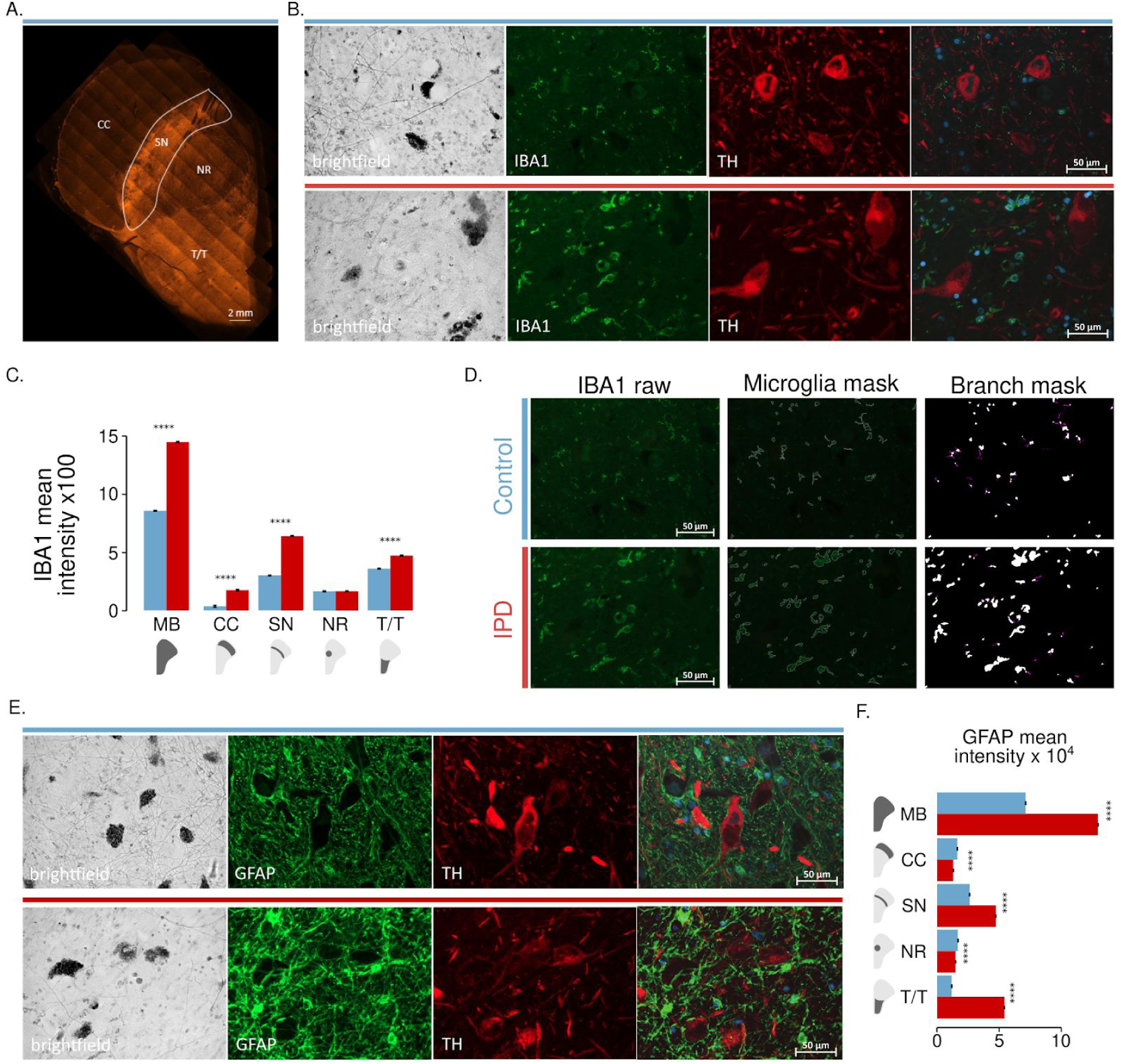
Quantitative immunofluorescence analysis of control and IPD midbrain microglia and astrocytes. (A) Midbrain regionalization for imaging highlighted in a MAP2 staining. (B) Representative control and IPD sections in brightfield (neuromelanin deposits) or stained for IBA1 and TH. (C) Microglia quantification. IBA1 intensity. (D) Microglia branching quantification strategy. IBA1 raw image, cell area segmentation, and branch masking. (E) Representative control and IPD sections in brightfield (neuromelanin deposits) or stained for GFAP and TH. Astrocyte quantification. (F) GFAP intensity. MB, midbrain; SN, substantia nigra; NR, nucleus ruber; T/T, tectum/tegmentum; and CC, crus cerebri. PD, red bar; control, blue bar; scale bar = 50μm; (* p < 0.1, ** p < 0.01, *** p < 0.001, **** p < 0.0001).

**Fig S3.**
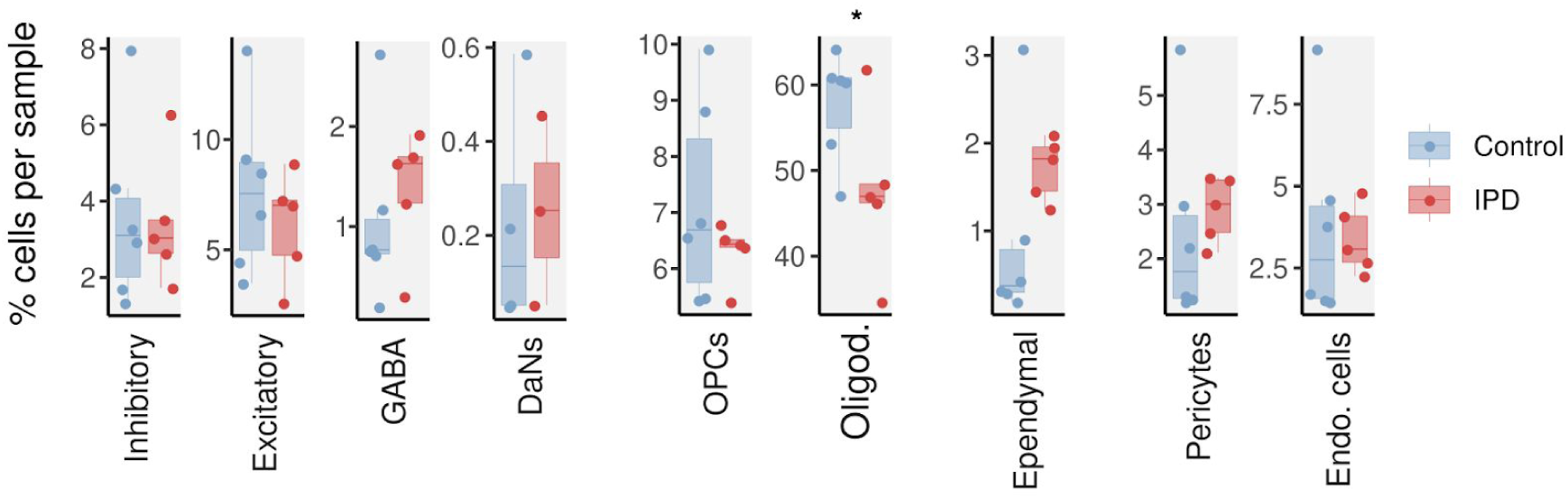
IPD differential cell type composition. Percentage of cells per sample is presented for control individuals and IPD patients. Inhibitory, excitatory, GABA and DaNs neurons, OPCs, oligodendrocytes (t test *p* = 0.08), ependymal and the vascular cells. (* p < 0.1, ** p < 0.01, *** p < 0.001, **** p < 0.0001).

**Fig S4.**
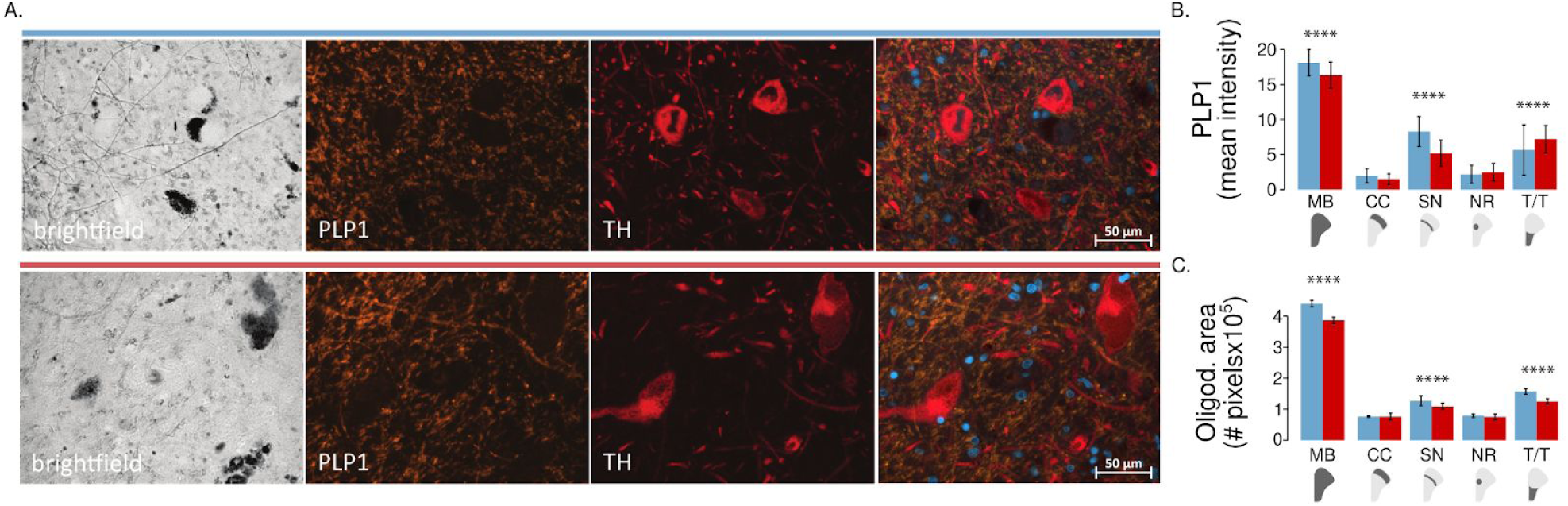
Quantitative immunofluorescence analysis of control and IPD midbrain oligodendrocytes. (A) Representative microscopy images of SN of control and IPD patient in brightfield (neuromelanin deposits), PLP1 (oligodendrocyte marker), TH. (B) PLP1 fluorescence intensity and (C) oligodendrocyte area. MB, midbrain; SN, substantia nigra; NR, nucleus ruber; T/T, tectum/tegmentum; and CC, crus cerebri. PD, red bar; control, blue bar; scale bar = 50μm; (* p < 0.1, ** p < 0.01, *** p < 0.001, **** p < 0.0001).

**Fig S5.**
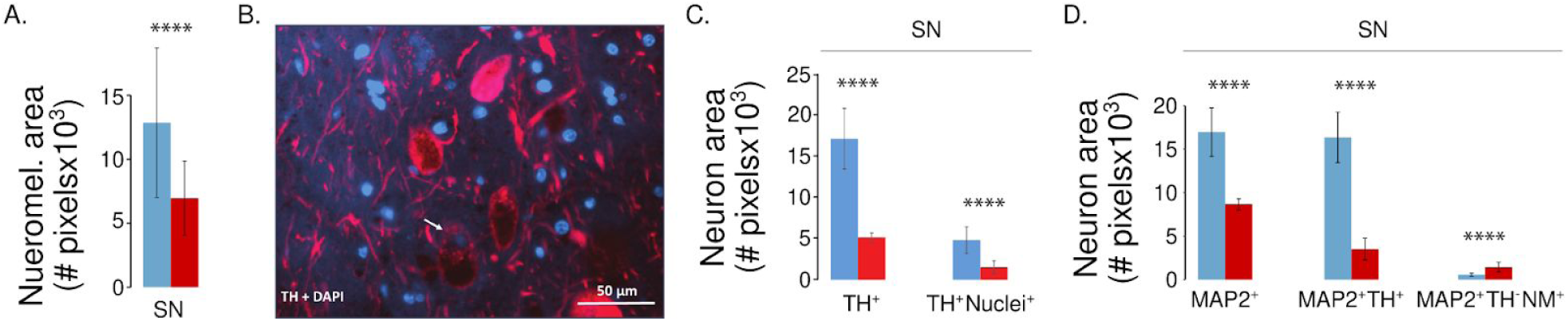
Quantitative immunofluorescence analysis of control and IPD midbrain neurons. (A) Neuromelanin content in a control individual and a IPD patient. (B) DaNs have large cell bodies of ∼50 μm width. As a consequence, individual nuclei may be “lost” during the sectioning at a thickness of 15 μm. (C) Neuronal TH-positive objects without nuclei (TH+) and such with nuclei (TH+/Nuclei+) per image frame. This analysis showed that only a subset of DaNs contains a nucleus in the individual sections. (D) Area of MAP2+/TH+/NM+ and MAP2+/TH-/NM+ neuronal cell populations in SN. While the area of MAP2+/TH+/NM+ neurons is reduced in IPD, MAP2+/TH-/NM+ neurons are more abundant in IPD than controls. SN, substantia nigra; PD, red bar; control, blue bar; scale bar = 50μm; (* p < 0.1, ** p < 0.01, *** p < 0.001, **** p < 0.0001).

**Fig S6.**
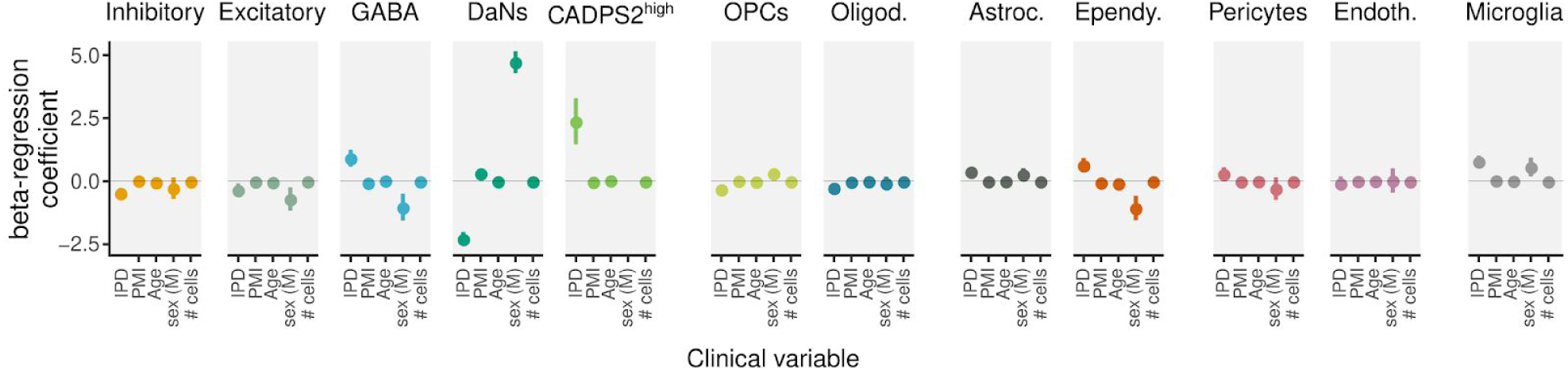
Cell type proportion beta-regression modeling estimates for the clinical variables: disease condition (PD), PMI, age, sex (M, male), and number of cells detected.

**Fig S7.**
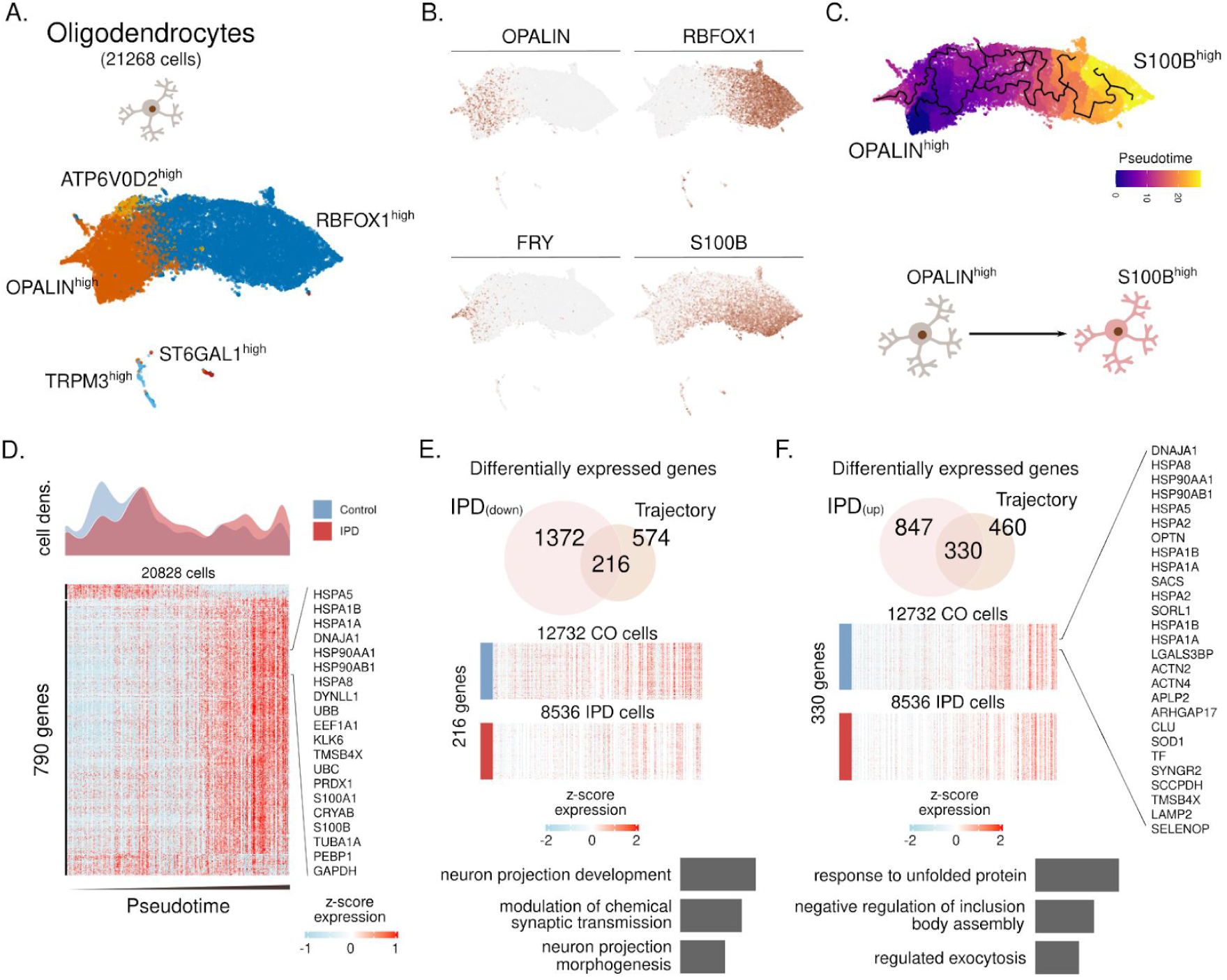
Trajectory inference reveals a loss of myelinating (OPALIN^high^) oligodendrocytes and differential activation in IPD. (A) Oligodendrocyte subpopulations named based on representative marker genes. (**B**) Expression of *OPALIN, RBFOX1, FRY* and *S100B* in the ∼21 thousand oligodendrocytes. (**C**) Inferred cell trajectory and pseudotime ordering of the major oligodendrocytes subpopulations, OPALIN^high^, ATP6V0D2^high^, and S100B^high^ cells. (**D**) IPD and control differential cell density over pseudotime. Expression levels of 790 highly variable genes across the oligodendrocyte trajectory. Expression is presented for ∼20 thousand oligodendrocytes organized by their pseudotime. (**E-F**) Intersection of IPD-differentially expressed and trajectory-associated genes. Also, the GO molecular enrichment of the intersected genes is presented. (**F**) 216 IPD-downregulated genes across the trajectory are associated with pathways important for neuron projection and synaptic transmission. (**F**) 330 genes are IPD-upregulated along the oligodendrocyte trajectory. These genes are mainly associated with the unfolded protein response.

## Supplementary tables

**Table S1.** Clinical and neuropathology features of control and IPD patients. PMI, post-mortem interval; SN, Substantia nigra.

| Patient | Sex | Age at onset (years) | Age at death (years) | PMI (h) | Lewy body Braak stage | Non-motor features | Profiled nuclei | SN neuron loss |
| --- | --- | --- | --- | --- | --- | --- | --- | --- |
| IPD1 | F | 70-75 | 80-85 | 25 | 5 | Depression, visual hallucinations | 1943 | severe |
| IPD2 | M | 55-60 | 65-70 | 24 | 6 | Hallucinations, paranoia | 5925 | severe |
| IPD3 | M | 70-75 | 75-80 | 24 | 6 | Stroke, cognitive impairment, dementia | 3345 | severe |
| IPD4 | M | 65-70 | 80-85 | 13 | 6 | Cognitive impairment, hallucinations, depression | 2083 | severe |
| IPD5 | M | 70-75 | 75-80 | 25 | 5 | Blindness, memory impairment | 5706 | severe |
| C1 | F | - | 90-95 | 29 | - | - | 3753 | mild |
| C2 | M | - | 65-70 | 16 | - | - | 4358 | none |
| C3 | M | - | 75-80 | 22 | - | - | 2047 | none |
| C4 | M | - | 85-90 | 5 | - | - | 4199 | none |
| C5 | M | - | 85-90 | 8 | - | - | 2318 | none |
| C6 | M | - | 85-90 | 12 | - | - | 5758 | none |

**Table S2.**
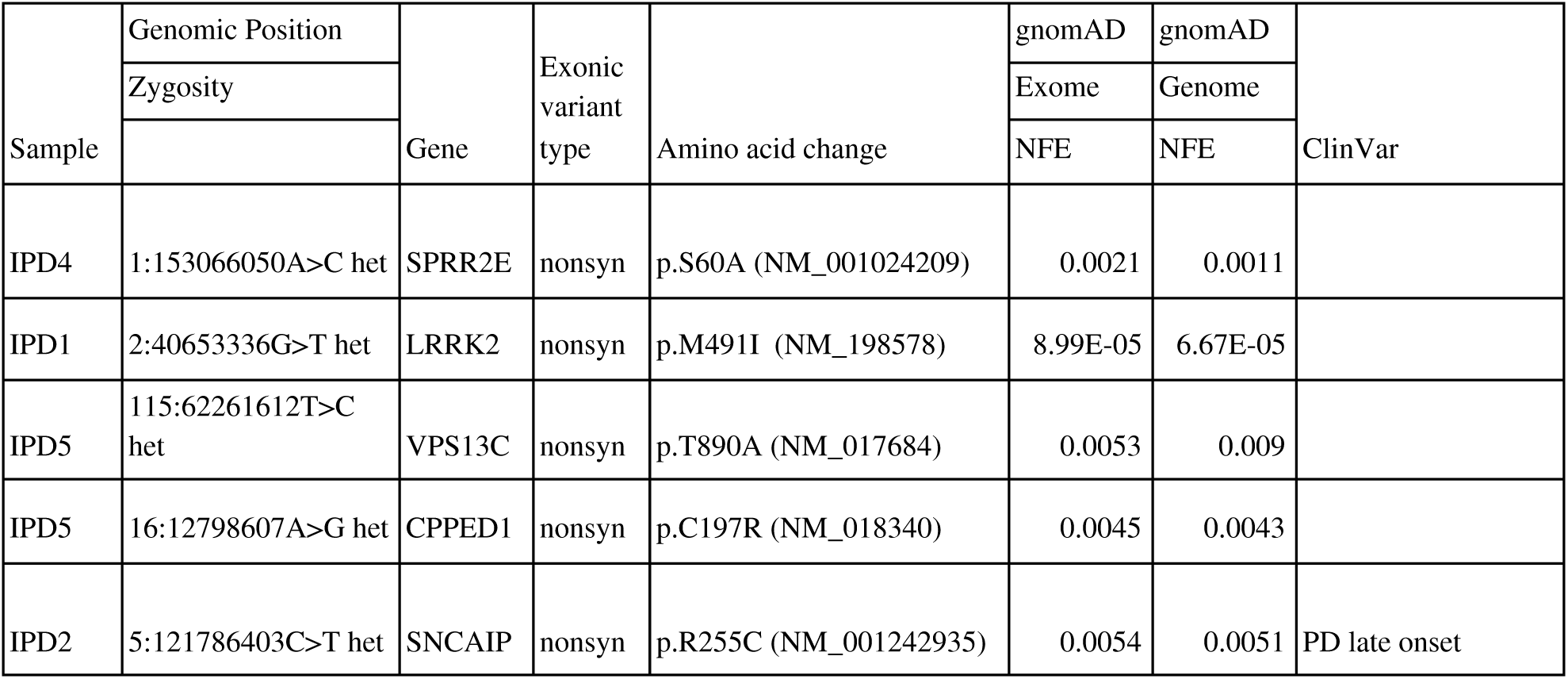
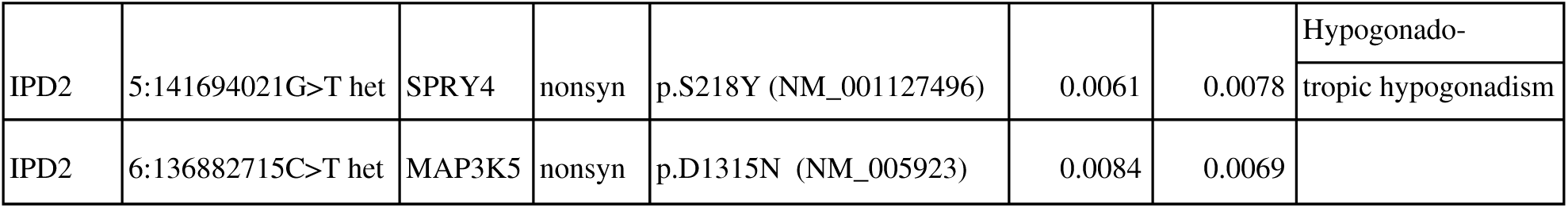
Genotyping results in IPD patients. gnomAD, Genome Aggregation Database and NFE, non-Finish European ethnicity for variant frequency in Exome and GnomAD database for NFE; het, heterozygous; nonsyn, nonsynonymous.

**Table S3.** Midbrain marker genes.

**Table S4.** PD-associated variant enrichment in genes with cell type-specific expression patterns. Ngenes, the number of genes; Beta, the regression coefficient of the variable; Beta SD, the semi-standardized regression coefficient, corresponding to the predicted change in Z-value given a change of one standard deviation in the predictor gene set; SE, the standard error of the regression coefficient; P, p-value for the variable.

| Cell type | Ngenes | Beta | Beta SD | SE | P |
| --- | --- | --- | --- | --- | --- |
| Astrocytes | 288 | 0.11462 | 0.014398 | 0.056003 | 0.020357 |
| CADPS2 | 135 | 0.059469 | 0.0051366 | 0.083315 | 0.23768 |
| DaNs | 3558 | 0.038082 | 0.015179 | 0.01739 | 0.014273 |
| Ependymal | 789 | 0.026188 | 0.0053671 | 0.033006 | 0.21377 |
| Excitatory | 3326 | 0.044136 | 0.017145 | 0.017837 | 0.006678 |
| GABA | 1944 | 0.014431 | 0.0044836 | 0.022251 | 0.25832 |
| Inhibitory | 3653 | 0.038517 | 0.015505 | 0.017311 | 0.013046 |
| Microglia | 312 | 0.13832 | 0.018072 | 0.050785 | 0.0032328 |
| ODC | 545 | 0.024718 | 0.0042402 | 0.039123 | 0.26376 |
| OPC | 346 | 0.098977 | 0.013605 | 0.053438 | 0.032011 |
| endothelial | 425 | -0.0094671 | -0.001439 | 0.045047 | 0.58323 |

**Table S5.** Per-gene information for every significant cell type within the MAGMA enrichment analysis. GENE, the gene ID; CHR, the chromosome the gene is on; START/STOP, the annotation boundaries of the gene on that chromosome (this includes an extended window of 35 kb upstream and 10 kb downstream of each gene); NSNPS, the number of SNPs annotated to that gene; NPARA, the number of relevant parameters used in the model (set to the number of principal components retained after pruning); N, the sample size used when analysing that gene; ZSTAT, the Z-value for the gene, based on its p-value; P, the gene p-value.

**Table S6.** Cell-type proportion differences. (t-test)

| cell_type | mean_%cell_control | mean_%cell_IPD | statistic | parameter | p.value |
| --- | --- | --- | --- | --- | --- |
| Astrocytes | 0.1003201348 | 0.1350897162 | -2.750440529 | 5.671063782 | 0.03527224809 |
| CADPS2+ neurons | 0.0006756662773 | 0.00716654498 | -4.143674419 | 3.08556866 | 0.02416713591 |
| DaNs | 0.002254081783 | 0.002534306896 | -0.1628600317 | 4.926478174 | 0.8770978439 |
| Endothelial cells | 0.03712579331 | 0.03379641715 | 0.2551487087 | 6.405729353 | 0.8066043354 |
| Ependymal | 0.008691805579 | 0.01714801578 | -1.763472461 | 6.174108097 | 0.1268821922 |
| Excitatory | 0.07705083796 | 0.06105230529 | 0.8389981942 | 8.563349355 | 0.4242711062 |
| GABA | 0.01057109988 | 0.01355919251 | -0.6523500725 | 8.899491317 | 0.5306619457 |
| Inhibitory | 0.03591306145 | 0.03436051229 | 0.1246382728 | 8.849982953 | 0.9035949956 |
| Microglia | 0.05494681848 | 0.1293926796 | -2.967776818 | 4.335692391 | 0.03727960072 |
| OPCs | 0.0717005494 | 0.06304498374 | 1.108974963 | 5.978794357 | 0.3100451593 |
| Oligodendrocytes | 0.5772245274 | 0.4762349461 | 2.004862556 | 6.698913849 | 0.08684078533 |
| Pericytes | 0.0247274284 | 0.0290674112 | -0.5552624256 | 6.296609446 | 0.5978755141 |

**Table S7.** T-test for microglia, astrocyte and oligodendrocyte staining.

| MICROGLIA |  |  |  |  |  |  |
| --- | --- | --- | --- | --- | --- | --- |
| Area | P value | Mean of Control | Mean of IPD | Difference | SE of difference | t ratio |
| CC | <0.000001 | 640.1 | 3277 | -2637 | 38.79 | 67.98 |
| NR | 0.011193 | 4455 | 4537 | -82.27 | 32.32 | 2.545 |
| TT | <0.000001 | 10038 | 10375 | -337.3 | 22.39 | 15.06 |
| SN | <0.000001 | 7772 | 43730 | -35958 | 439 | 81.92 |
| Midbrain | <0.000001 | 22905 | 61919 | -39014 | 182.8 | 213.5 |
| Intensity | P value | Mean of Control | Mean of IPD | Difference | SE of difference | t ratio |
| CC | <0.000001 | 34.4 | 172.7 | -138.3 | 1.011 | 136.7 |
| NR | 0.277261 | 163.9 | 164.3 | -0.4015 | 0.3692 | 1.088 |
| TT | <0.000001 | 357.3 | 469.6 | -112.3 | 0.2625 | 427.7 |
| SN | <0.000001 | 298.5 | 637.4 | -338.9 | 0.2215 | 1530 |
| Midbrain | <0.000001 | 854.2 | 1444 | -589.8 | 0.1693 | 3484 |
| ASTROCYTES |  |  |  |  |  |  |
| Area | P value | Mean of Control | Mean of IPD | Difference | SE of difference | t ratio |
| CC | <0.000001 | 7928 | 5717 | 2211 | 90.19 | 24.51 |
| NR | <0.000001 | 149673 | 198551 | -48878 | 30.12 | 1623 |
| TT | <0.000001 | 52452 | 258069 | -205617 | 24.76 | 8303 |
| SN | <0.000001 | 126058 | 198274 | -72216 | 26.02 | 2776 |
| Midbrain | <0.000001 | 336111 | 660610 | -324500 | 73.43 | 4419 |
| Intensity | P value | Mean of Control | Mean of IPD | Difference | SE of difference | t ratio |
| CC | <0.000001 | 16389 | 13030 | 3359 | 0.5952 | 5644 |
| NR | <0.000001 | 16747 | 14928 | 1819 | 0.1765 | 10307 |

| TT | <0.000001 | 11802 | 54248 | -42446 | 0.1493 | 284322 |
| --- | --- | --- | --- | --- | --- | --- |
| SN | <0.000001 | 26300 | 47340 | -21040 | 0.1498 | 140471 |
| Midbrain | <0.000001 | 71238 | 129546 | -58308 | 0.07306 | 798099 |
| OLIGODENDROCYTES |  |  |  |  |  |  |
| Area | P value | Mean of Control | Mean of Midbrain | Difference | SE of difference | t ratio |
| CC | 0.931916 | 76517 | 76257 | 260.5 | 3007 | 0.08664 |
| NR | 0.152874 | 79237 | 75183 | 4054 | 2791 | 1.452 |
| TT | <0.000001 | 157022 | 125198 | 31824 | 1745 | 18.24 |
| SN | <0.000001 | 127404 | 109638 | 17767 | 517.3 | 34.34 |
| Midbrain | <0.000001 | 440181 | 386276 | 53905 | 424.2 | 127.1 |
| Intensity | P value | Mean of Control | Mean of Midbrain | Difference | SE of difference | t ratio |
| CC | 0.28854 | 1.971 | 1.482 | 0.4889 | 0.447 | 1.094 |
| NR | 0.494075 | 2.176 | 2.459 | -0.2832 | 0.411 | 0.6891 |
| TT | 0.00003 | 5.674 | 7.206 | -1.532 | 0.3644 | 4.204 |
| SN | <0.000001 | 8.295 | 5.183 | 3.112 | 0.08408 | 37.01 |
| Midbrain | <0.000001 | 18.12 | 16.33 | 1.786 | 0.07642 | 23.37 |

**Table S8.**
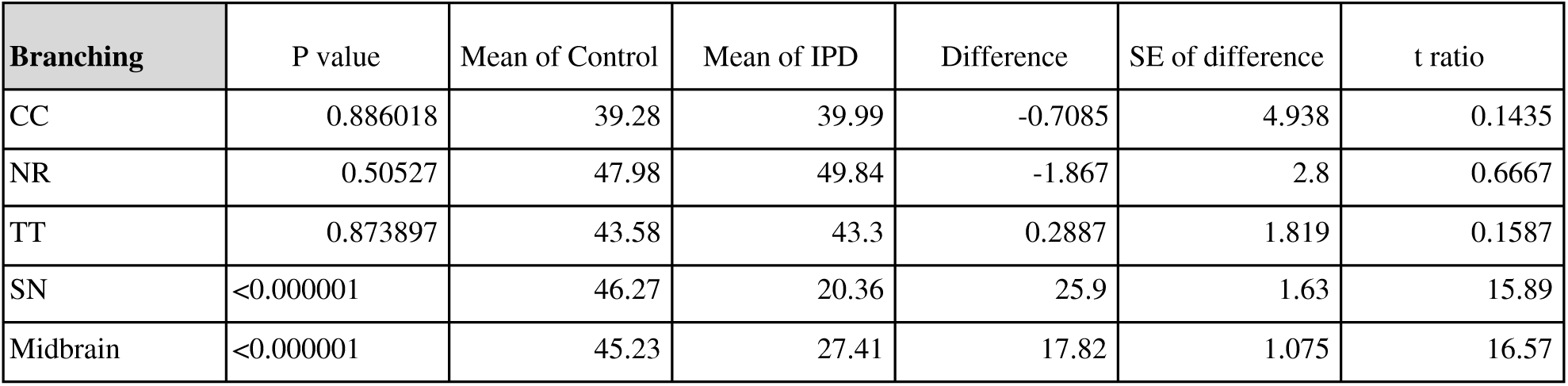
T-test for microglia branching.

| Branching | P value | Mean of Control | Mean of IPD | Difference | SE of difference | t ratio |
| --- | --- | --- | --- | --- | --- | --- |
| CC | 0.886018 | 39.28 | 39.99 | -0.7085 | 4.938 | 0.1435 |
| NR | 0.50527 | 47.98 | 49.84 | -1.867 | 2.8 | 0.6667 |
| TT | 0.873897 | 43.58 | 43.3 | 0.2887 | 1.819 | 0.1587 |
| SN | <0.000001 | 46.27 | 20.36 | 25.9 | 1.63 | 15.89 |
| Midbrain | <0.000001 | 45.23 | 27.41 | 17.82 | 1.075 | 16.57 |

**Table S9.**
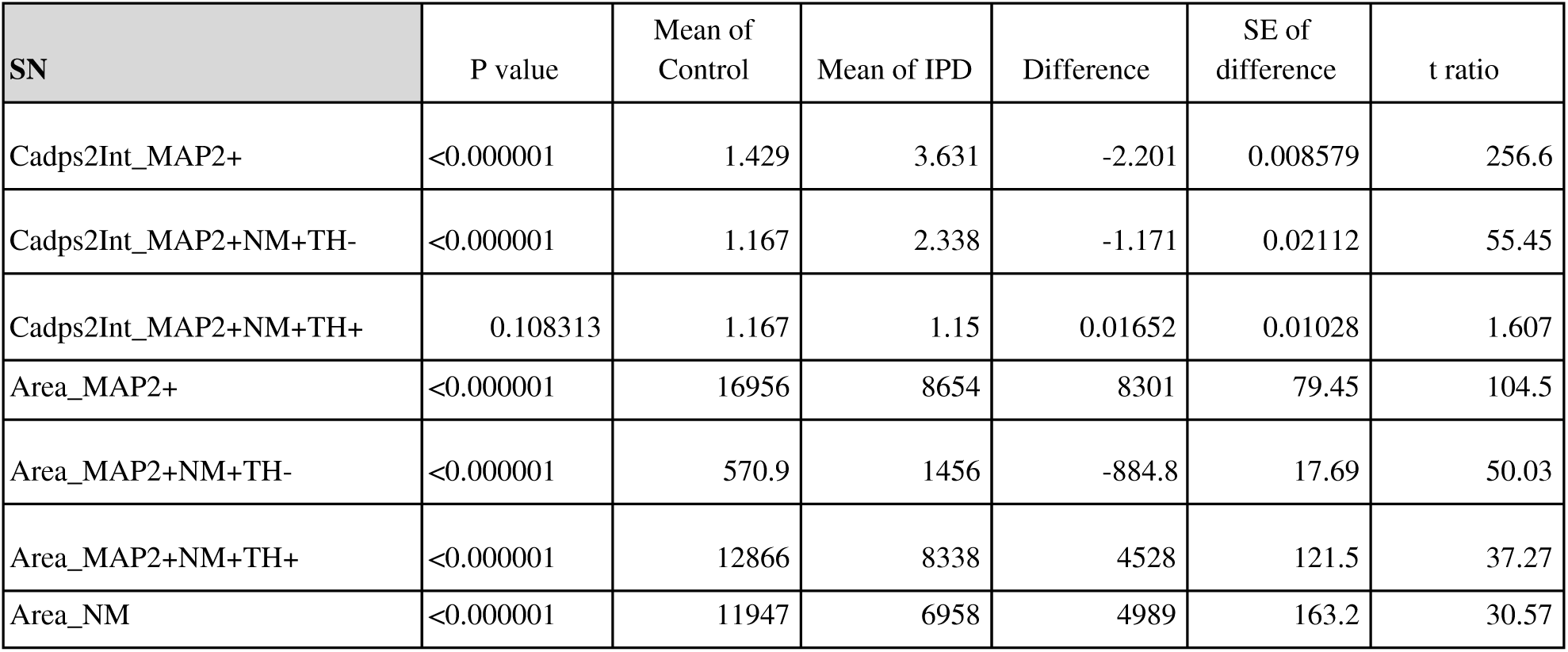
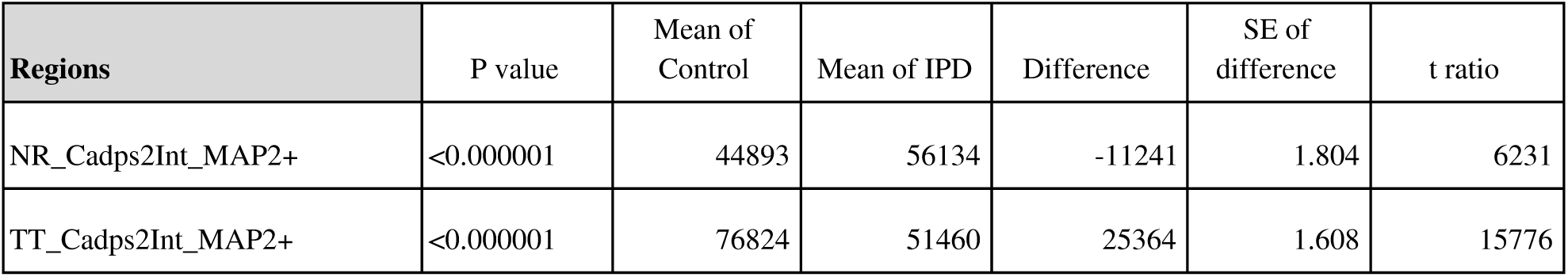
T-test for neuronal staining.

## References and Notes

1. Grünewald, A., Kumar, K. R. & Sue, C. M. New insights into the complex role of mitochondria in Parkinson’s disease. Prog. Neurobiol. 177, 73–93 (2019).

2. Smolders, S. & Van Broeckhoven, C. Genetic perspective on the synergistic connection between vesicular transport, lysosomal and mitochondrial pathways associated with Parkinson’s disease pathogenesis. Acta Neuropathol Commun 8, 63 (2020).

3. Inamdar, N. N., Arulmozhi, D. K., Tandon, A. & Bodhankar, S. L. Parkinson’s disease: genetics and beyond. Curr. Neuropharmacol. 5, 99–113 (2007).

4. Obeso, J. A. et al. Past, present, and future of Parkinson’s disease: A special essay on the 200th Anniversary of the Shaking Palsy. Mov. Disord. 32, 1264–1310 (2017).

5. Armstrong, M. J. & Okun, M. S. Diagnosis and Treatment of Parkinson Disease: A Review. JAMA 323, 548–560 (2020).

6. Blesa, J. & Przedborski, S. Parkinson’s disease: animal models and dopaminergic cell vulnerability. Front. Neuroanat. 8, 155 (2014).

7. Borrageiro, G., Haylett, W., Seedat, S., Kuivaniemi, H. & Bardien, S. A review of genome-wide transcriptomics studies in Parkinson’s disease. Eur. J. Neurosci. 47, 1–16 (2018).

8. Cao, J. et al. The single-cell transcriptional landscape of mammalian organogenesis. Nature 566, 496–502 (2019).

9. Blauwendraat, C. et al. NeuroChip, an updated version of the NeuroX genotyping platform to rapidly screen for variants associated with neurological diseases. Neurobiol. Aging 57, 247.e9–247.e13 (2017).

10. Mitkus, S. N. et al. Expression of oligodendrocyte-associated genes in dorsolateral prefrontal cortex of patients with schizophrenia. Schizophr. Res. 98, 129–138 (2008).

11. van Bruggen, D., Agirre, E. & Castelo-Branco, G. Single-cell transcriptomic analysis of oligodendrocyte lineage cells. Curr. Opin. Neurobiol. 47, 168–175 (2017).

12. Ikeshima-Kataoka, H. Neuroimmunological Implications of AQP4 in Astrocytes. Int. J. Mol. Sci. 17, (2016).

13. Shah, P. T. et al. Single-Cell Transcriptomics and Fate Mapping of Ependymal Cells Reveals an Absence of Neural Stem Cell Function. Cell 173, 1045–1057.e9 (2018).

14. Hwang, I. K. et al. CD74-immunoreactive activated M1 microglia are shown late in the gerbil hippocampal CA1 region following transient cerebral ischemia. Mol. Med. Rep. 15, 4148–4154 (2017).

15. Maher, T. J. et al. ATP-binding cassette transporter Abcg2 lineage contributes to the cardiac vasculature after oxidative stress. Am. J. Physiol. Heart Circ. Physiol. 306, H1610–8 (2014).

16. Jang, A. S. et al. Endothelial dysfunction and claudin 5 regulation during acrolein-induced lung injury. Am. J. Respir. Cell Mol. Biol. 44, 483–490 (2011).

17. Bell, R. D. et al. Pericytes control key neurovascular functions and neuronal phenotype in the adult brain and during brain aging. Neuron 68, 409–427 (2010).

18. Kodama, T. et al. Neuronal classification and marker gene identification via single-cell expression profiling of brainstem vestibular neurons subserving cerebellar learning. J. Neurosci. 32, 7819–7831 (2012).

19. Merrill, C. B., Friend, L. N., Newton, S. T., Hopkins, Z. H. & Edwards, J. G. Ventral tegmental area dopamine and GABA neurons: Physiological properties and expression of mRNA for endocannabinoid biosynthetic elements. Sci. Rep. 5, 16176 (2015).

20. Wu, L.-J. et al. Increased anxiety-like behavior and enhanced synaptic efficacy in the amygdala of GluR5 knockout mice. PLoS One 2, e167 (2007).

21. Thompson, L., Barraud, P., Andersson, E., Kirik, D. & Björklund, A. Identification of dopaminergic neurons of nigral and ventral tegmental area subtypes in grafts of fetal ventral mesencephalon based on cell morphology, protein expression, and efferent projections. J. Neurosci. 25, 6467–6477 (2005).

22. Spillantini, M. G. et al. α-Synuclein in Lewy bodies. Nature vol. 388 839–840 (1997).

23. Schapansky, J., Nardozzi, J. D. & LaVoie, M. J. The complex relationships between microglia, alpha-synuclein, and LRRK2 in Parkinson’s disease. Neuroscience 302, 74–88 (2015).

24. Grünewald, A. et al. Quantitative quadruple-label immunofluorescence of mitochondrial and cytoplasmic proteins in single neurons from human midbrain tissue. J. Neurosci. Methods 232, 143–149 (2014).

25. Torres-Platas, S. G. et al. Morphometric characterization of microglial phenotypes in human cerebral cortex. J. Neuroinflammation 11, 12 (2014).

26. Walker, D. G. et al. Patterns of Expression of Purinergic Receptor P2RY12, a Putative Marker for Non-Activated Microglia, in Aged and Alzheimer’s Disease Brains. Int. J. Mol. Sci. 21, (2020).

27. van der Poel, M. et al. Transcriptional profiling of human microglia reveals grey-white matter heterogeneity and multiple sclerosis-associated changes. Nat. Commun. 10, 1139 (2019).

28. Kakimura, J.-I. et al. Microglial activation and amyloid-beta clearance induced by exogenous heat-shock proteins. FASEB J. 16, 601–603 (2002).

29. Burm, S. M. et al. Inflammasome-induced IL-1β secretion in microglia is characterized by delayed kinetics and is only partially dependent on inflammatory caspases. J. Neurosci. 35, 678–687 (2015).

30. Hüttenrauch, M. et al. Glycoprotein NMB: a novel Alzheimer’s disease associated marker expressed in a subset of activated microglia. Acta Neuropathol Commun 6, 108 (2018).

31. Parajuli, B. et al. Oligomeric amyloid β induces IL-1β processing via production of ROS: implication in Alzheimer’s disease. Cell Death Dis. 4, e975 (2013).

32. Nalls, M. A. et al. Large-scale meta-analysis of genome-wide association data identifies six new risk loci for Parkinson’s disease. Nat. Genet. 46, 989–993 (2014).

33. Wu, K.-C., Liou, H.-H., Kao, Y.-H., Lee, C.-Y. & Lin, C.-J. The critical role of Nramp1 in degrading α-synuclein oligomers in microglia under iron overload condition. Neurobiol. Dis. 104, 61–72 (2017).

34. Li, S. et al. The accelerated aging model reveals critical mechanisms of late-onset Parkinson’s disease. BioData Min. 13, 4 (2020).

35. Bradford, B. M., Wijaya, C. A. W. & Mabbott, N. A. Discrimination of Prion Strain Targeting in the Central Nervous System via Reactive Astrocyte Heterogeneity in CD44 Expression. Front. Cell. Neurosci. 13, 411 (2019).

36. Smith, H. L. et al. Astrocyte Unfolded Protein Response Induces a Specific Reactivity State that Causes Non-Cell-Autonomous Neuronal Degeneration. Neuron 105, 855–866.e5 (2020).

37. Muchowski, P. J. & Wacker, J. L. Modulation of neurodegeneration by molecular chaperones. Nat. Rev. Neurosci. 6, 11–22 (2005).

38. Jiang, W. et al. Identification of Tmem10 as a novel late-stage oligodendrocytes marker for detecting hypomyelination. Int. J. Biol. Sci. 10, 33–42 (2013).

39. Sathe, K. et al. S100B is increased in Parkinson’s disease and ablation protects against MPTP-induced toxicity through the RAGE and TNF-α pathway. Brain 135, 3336–3347 (2012).

40. Čajánek, L. et al. Tiam1 regulates the Wnt/Dvl/Rac1 signaling pathway and the differentiation of midbrain dopaminergic neurons. Mol. Cell. Biol. 33, 59–70 (2013).

41. Höglinger, G. U. et al. The pRb/E2F cell-cycle pathway mediates cell death in Parkinson’s disease. Proc. Natl. Acad. Sci. U. S. A. 104, 3585–3590 (2007).

42. Joers, V., Tansey, M. G., Mulas, G. & Carta, A. R. Microglial phenotypes in Parkinson’s disease and animal models of the disease. Prog. Neurobiol. 155, 57–75 (2017).

43. Alessi, D. R. & Sammler, E. LRRK2 kinase in Parkinson’s disease. Science 360, 36–37 (2018).

44. Smith, J. A. Regulation of Cytokine Production by the Unfolded Protein Response; Implications for Infection and Autoimmunity. Front. Immunol. 9, 422 (2018).

45. Dong, Y. & Benveniste, E. N. Immune function of astrocytes. Glia 36, 180–190 (2001).

46. Hamanaka, G., Ohtomo, R., Takase, H., Lok, J. & Arai, K. White-matter repair: Interaction between oligodendrocytes and the neurovascular unit. Brain Circ 4, 118–123 (2018).

47. Nalls, M. A. et al. Identification of novel risk loci, causal insights, and heritable risk for Parkinson’s disease: a meta-analysis of genome-wide association studies. Lancet Neurol. 18, 1091–1102 (2019).

48. Deloulme, J. C. et al. Nuclear expression of S100B in oligodendrocyte progenitor cells correlates with differentiation toward the oligodendroglial lineage and modulates oligodendrocytes maturation. Mol. Cell. Neurosci. 27, 453–465 (2004).

49. Sheng, J. G. et al. In vivo and in vitro evidence supporting a role for the inflammatory cytokine interleukin-1 as a driving force in Alzheimer pathogenesis. Neurobiol. Aging 17, 761–766 (1996).

50. Smits, L. M. et al. Single-cell transcriptomics reveals multiple neuronal cell types in human midbrain-specific organoids. 589598 (2019) doi:10.1101/589598.

51. Grima, B. et al. A single human gene encoding multiple tyrosine hydroxylases with different predicted functional characteristics. Nature 326, 707–711 (1987).

52. Tsuda, L. & Lim, Y.-M. Regulatory system for the G1-arrest during neuronal development in Drosophila. Dev. Growth Differ. 56, 358–367 (2014).

53. Brunk, I., Blex, C., Speidel, D., Brose, N. & Ahnert-Hilger, G. Ca2+-dependent activator proteins of secretion promote vesicular monoamine uptake. J. Biol. Chem. 284, 1050–1056 (2009).

54. Reinhardt, P. et al. Genetic correction of a LRRK2 mutation in human iPSCs links parkinsonian neurodegeneration to ERK-dependent changes in gene expression. Cell Stem Cell 12, 354–367 (2013).

55. Arenas, E. Wnt signaling in midbrain dopaminergic neuron development and regenerative medicine for Parkinson’s disease. J. Mol. Cell Biol. 6, 42–53 (2014).

56. Carballo-Carbajal, I. et al. Brain tyrosinase overexpression implicates age-dependent neuromelanin production in Parkinson’s disease pathogenesis. Nat. Commun. 10, 973 (2019).

57. Wolock, S. L., Lopez, R. & Klein, A. M. Scrublet: Computational Identification of Cell Doublets in Single-Cell Transcriptomic Data. Cell Syst 8, 281–291.e9 (2019).

58. Stuart, T. et al. Comprehensive Integration of Single-Cell Data. Cell 177, 1888–1902.e21 (2019).

59. Hafemeister, C. & Satija, R. Normalization and variance stabilization of single-cell RNA-seq data using regularized negative binomial regression. Genome Biol. 20, 296 (2019).

60. McInnes, L., Healy, J. & Melville, J. UMAP: Uniform Manifold Approximation and Projection for Dimension Reduction. arXiv [stat.ML] (2018).

61. Modern Applied Statistics with S, 4th ed. https://www.stats.ox.ac.uk/pub/MASS4/.

62. R. D. C. Team. R: A language and environment for statistical computing (ISBN 3-900051-07-0). https://www.scienceopen.com/document?vid=300a2dc0-3207-4383-818c-51eb0f49f561.

63. Cribari-Neto, F. & Zeileis, A. Beta Regression in R. Journal of Statistical Software, Articles 34, 1–24 (2010).

64. Haghverdi, L., Lun, A. T. L., Morgan, M. D. & Marioni, J. C. Batch effects in single-cell RNA-sequencing data are corrected by matching mutual nearest neighbors. Nat. Biotechnol. 36, 421–427 (2018).

65. Chen, E. Y. et al. Enrichr: interactive and collaborative HTML5 gene list enrichment analysis tool. BMC Bioinformatics 14, 128 (2013).

66. Maher, M. P., Pine, J., Wright, J. & Tai, Y. C. The neurochip: a new multielectrode device for stimulating and recording from cultured neurons. J. Neurosci. Methods 87, 45–56 (1999).

67. Reed, E. et al. A guide to genome-wide association analysis and post-analytic interrogation. Stat. Med. 34, 3769–3792 (2015).

68. Das, S. et al. Next-generation genotype imputation service and methods. Nat. Genet. 48, 1284–1287 (2016).

69. de Leeuw, C. A., Mooij, J. M., Heskes, T. & Posthuma, D. MAGMA: generalized gene-set analysis of GWAS data. PLoS Comput. Biol. 11, e1004219 (2015).

70. Reynolds, R. H. et al. Moving beyond neurons: the role of cell type-specific gene regulation in Parkinson’s disease heritability. NPJ Parkinsons Dis 5, 6 (2019).

71. Korsunsky, I. et al. Fast, sensitive and accurate integration of single-cell data with Harmony. Nat. Methods 16, 1289–1296 (2019).

72. Cleveland, W. S. Robust Locally Weighted Regression and Smoothing Scatterplots. null 74, 829–836 (1979).

73. Pedregosa, F. et al. Scikit-learn: Machine Learning in Python. J. Mach. Learn. Res. 12, 2825–2830 (2011).

74. Szymański, P., Kajdanowicz, T. & Kersting, K. How Is a Data-Driven Approach Better than Random Choice in Label Space Division for Multi-Label Classification? Entropy 18, 282 (2016).

75. Hagberg, A., Swart, P. & S Chult, D. Exploring network structure, dynamics, and function using networkx. https://www.osti.gov/biblio/960616-exploring-network-structure-dynamics-function-using-networkx (2008).

76. Blondel, V. D., Guillaume, J.-L., Lambiotte, R. & Lefebvre, E. Fast unfolding of communities in large networks. J. Stat. Mech. 2008, P10008 (2008).

